# Circulating gelsolin tracks disease stage in amyotrophic lateral sclerosis across four cohorts

**DOI:** 10.64898/2026.11.18.26368465

**Authors:** Andrew Strange, Marcel Weinreich, Hadar Klimovski, Cory Holdom, Alex Daniel, Mays Baidoun, Farah Mahiddine, Kari Wong, Iddo Magen, Yurui Chen, Martin Turner, Kevin Talbot, David G. Lester, Alexander G. Thompson, Eran Hornstein, Sai Zhang, Johnathan Cooper-Knock, Pamela J. Shaw

## Abstract

Amyotrophic lateral sclerosis is a fatal neurodegenerative disease with highly variable survival, and blood biomarkers beyond neurofilament light chain (NfL) are needed for prognosis and trial stratification. A biomarker of progress through an individualised disease course may be prognostic but independent of the rate of neurodegeneration.

We analysed the serum proteome (5,401 proteins) and plasma and cerebrospinal fluid metabolomes (1,659 metabolites) of 94 patients with amyotrophic lateral sclerosis and 39 controls recruited in Sheffield, UK, and replicated findings in cohorts from Tel Aviv, Oxford and the United States (AnswerALS). We defined “disease stage” as the fraction of the onset-to-death interval elapsed at sampling. In contrast we define the rate of neurodegeneration by total survival time or the rate of symptom progression quantified using the D50 model of functional decline.

Survival-associated proteins were enriched for stage associations in Sheffield (odds ratio = 5.1) and in the Oxford (2.9), and Tel Aviv (1.6) replication cohorts. Sixty-eight proteins not including NfL tracked stage across four cohorts, implicating systemic inflammation and muscle remodelling; the same biology is tracked by 149 plasma and 58 CSF stage-associated metabolites. Stage-associated proteins that rose with stage also rose with age in UK Biobank (17 of 17 age-associated proteins), and the proteomic age gap rose nominally with disease stage (partial Spearman r = 0.25, P = 0.027; BH-adjusted P = 0.055). Circulating gelsolin (GSN) fell with advancing disease in all four cohorts. NfL tracked disease aggressiveness (standardised β 0.56, P = 1.1 × 10⁻¹⁴) but not stage (0.13, P = 0.056), whereas GSN tracked stage (−0.41, P = 8.4 × 10⁻⁹) but not the rate of symptom progression (−0.08, P = 0.25). GSN explained 7–10% of the variance between patients in disease stage quantified by survival time and 19% of the variance in stage quantified by functional decline; this is comparable to the variance in the rate of neurodegeneration which is captured by circulating NfL.

Accounting for disease stage separates markers of disease rate from markers of disease position, refining both biomarker discovery and biological interpretation in amyotrophic lateral sclerosis (ALS).

## Introduction

Amyotrophic lateral sclerosis (ALS) is a fatal neurodegenerative disease of upper and lower motor neurons.^1^ Survival from symptom onset ranges from months to more than a decade,^2^ and this heterogeneity complicates both prognosis for individual patients and the design of clinical trials. Neurofilament light chain (NfL), a marker of the rate of neuroaxonal injury, remains the only well-established blood prognostic biomarker.^3,4^ An under-addressed aspect of prognostic biomarker discovery is that patients are sampled at different points in disease courses of very different lengths. Relative positioning within each patient’s disease course rather than absolute time elapsed carries important prognostic information; patients sampled later in their disease course are likely to die sooner. Framed in terms of symptom progression, this idea underpins the King’s clinical staging system which considers spread through the CNS,^5^ and the D50 model which calculates the time taken to lose half of functional capacity as measured using longitudinal ALSFRS-R scores.^6^ A circulating biomarker could be prognostic because it reflects how far the disease has progressed rather than how fast it is progressing. Separating these two components is important for biomarker discovery, for interpreting disease biology and for stratifying patients in trials.

High-throughput proteomics has recently been applied to ALS at scale. Chia et al. profiled 2,868 plasma proteins in 183 patients and 309 controls and identified 33 differentially abundant proteins.^7^ These proteins formed a diagnostic model and, in pre-symptomatic samples, pointed to involvement of skeletal muscle, nerve and energy metabolism years before onset. Ran et al. followed carriers of ALS-associated pathogenic variants longitudinally and identified 92 proteins that changed before phenoconversion, including NfL, EDA2R and CA3.^8^ A 19-protein panel predicted phenoconversion over 0.5 to 5 years. Two recent studies identified serum proteins associated with survival in large cohorts profiled across complementary platforms, several of which added prognostic information beyond NfL.^9,10^ In these studies, the stage of disease at which patients were sampled was not explicitly considered.

The circulating metabolome has also been linked to ALS risk and progression. Using Mendelian randomisation, we causally associated five metabolites with ALS risk.^11^ In directly measured patient biofluids, acetylcarnitine was associated with later onset, CSF lactate with longer survival and plasma fructose with shorter survival. Guo et al. related longitudinal plasma metabolomes to functional decline and found that lipid metabolites dominated, particularly fatty acids, sphingomyelins, plasmalogens and phosphatidylcholines.^12^ In the multi-omic METABALS cohort, however, untargeted metabolomic profiles added little prognostic value beyond serum NfL.^13^ Few studies have combined proteome, plasma and CSF metabolomes and survival in the same individuals.

Age is the strongest risk factor for ALS, and biological age has been estimated from molecular ageing clocks. Epigenetic age acceleration has been associated with earlier onset and shorter survival in ALS.^14^ In 428 patients, GrimAge acceleration was 1.8 years greater than in controls and, in men, was associated with shorter survival.^15^ In more than 5,000 patients from Project MinE, however, accelerated epigenetic ageing was explained by white blood cell composition, except in *C9orf72* expansion carriers, and did not predict survival.^16^ Proteomic clocks offer an alternative measure that is independent of cell composition. Argentieri et al. trained a 204-protein clock in 45,441 UK Biobank participants (r = 0.94 with chronological age).^17^ The resulting proteomic age gap predicted mortality, multimorbidity and incident neurodegenerative disease, and replicated in Chinese and Finnish biobanks. To our knowledge, proteomic ageing has not been examined in relation to the course of ALS.

Here we analysed the serum proteome and the plasma and CSF metabolomes of patients with ALS from the Sheffield arm of the AMBRoSIA study, with replication in cohorts from Tel Aviv, Oxford and the United States (AnswerALS).^18^ We defined disease stage as the fraction of each patient’s onset-to-death interval that had elapsed at sampling. In Oxford, we separately estimated disease aggressiveness and disease phase using the D50 model of functional decline.^6^ We show that circulating survival-associated proteins are strongly enriched for stage associations across cohorts, with the exception of NfL. Sixty-eight proteins track stage across four cohorts and implicate systemic inflammation and muscle remodelling. These stage-associated proteins mark disease phase rather than aggressiveness, overlap with proteomic ageing, and are accompanied by a nominal rise in proteomic age gap with advancing disease. Gelsolin (GSN), the most consistent stage-associated protein, marks disease stage and NfL the rate of decline, and together they improve survival prediction. Our approach is summarised in **Figure 1**.

**Figure 1.**
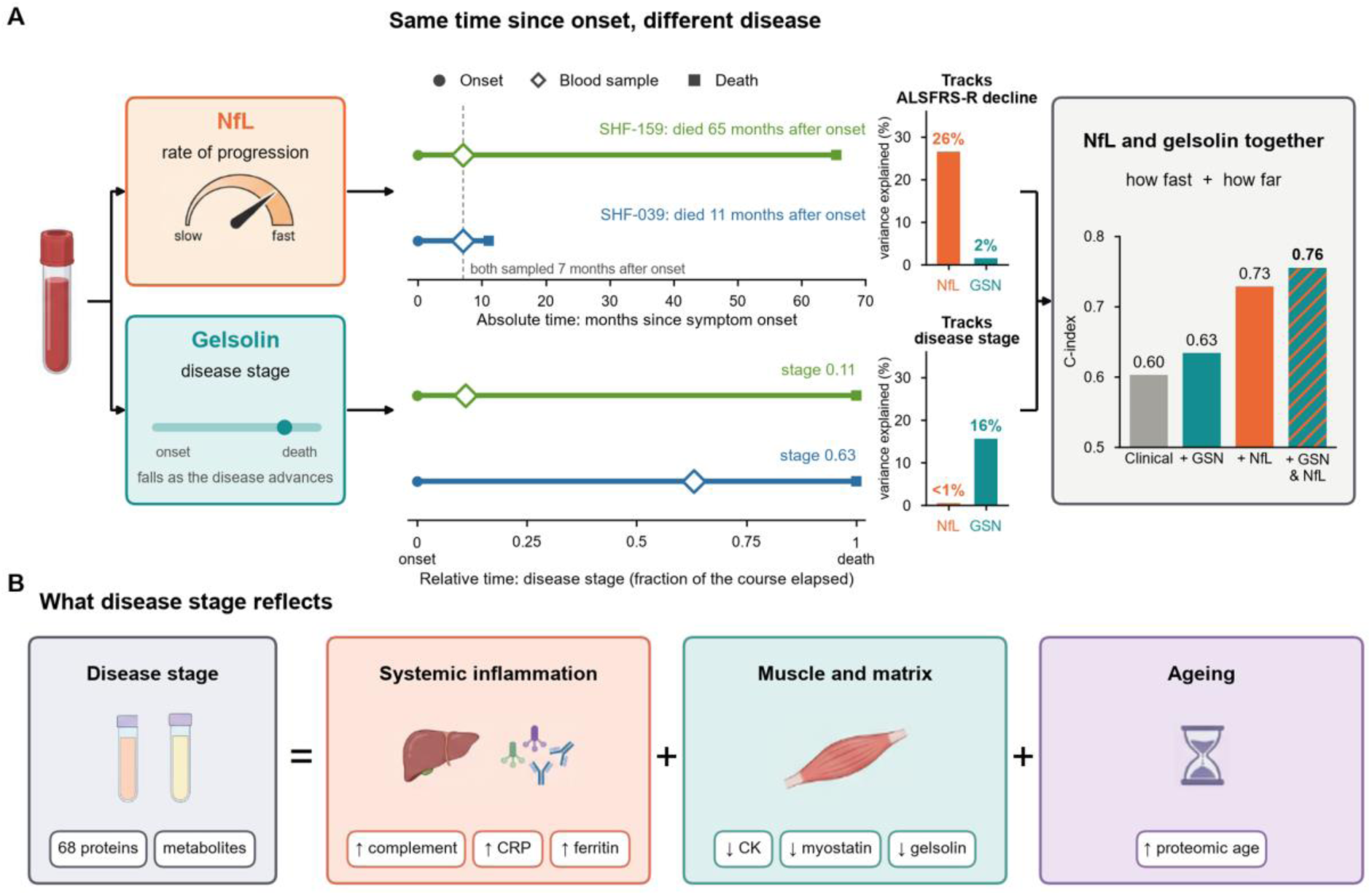
Disease rate, disease stage and the biology of stage. (**A**) *Left*: NfL (orange) tracks the rate of progression and gelsolin (teal) disease stage. *Middle*: Two example Sheffield patients illustrate these different measures of progression; both patients are sampled 7 months after symptom onset (absolute time, months since onset) but at very different disease stages (fraction of the onset-to-death interval elapsed at sampling; 0, onset; 1, death). SHF-159 (green) died after 65 months and SHF-039 (blue) died 11 months after onset; therefore they were sampled at stage 0.11 and 0.63. Filled circle, onset; open diamond, blood sample; square, death. Bars, variance in ALSFRS-R decline and in disease stage explained by each marker in Oxford (221 deceased participants; partial Spearman ρ², adjusted for age, sex, centre and the other dimension). *Right*: cross-validated C-index for survival from sampling in Sheffield and Oxford pooled (475 participants, 336 deaths; Cox models stratified by dataset; 5 × 10-fold cross-validation) with age and sex (clinical; grey) and with gelsolin, NfL or both (striped) added. (**B**) Schematic of the biology shared by stage-associated serum proteins and plasma and CSF metabolites: systemic inflammation (higher complement, C-reactive protein and ferritin), muscle and matrix (lower creatine kinase, myostatin and gelsolin) and ageing (higher proteomic age). Icons created in BioRender. ALSFRS-R, revised ALS Functional Rating Scale; CSF, cerebrospinal fluid; NfL, neurofilament light chain.

## Results

### ALS study cohorts and multi-omic profiling

Sheffield samples were collected at the baseline visit of the AMBRoSIA study (**Methods**). The serum proteome (Olink Explore HT) covers 142 participants (118 with MND, of whom 94 have ALS, and 24 controls) and 5,401 assays; the plasma metabolome (Metabolon HD4) covers 134 participants (95 MND, 39 controls) and 1,659 metabolites (1,304 named); and the CSF metabolome covers 110 participants (76 MND, 34 controls) and 513 metabolites (450 named) (**Table 1** and **Supplementary Table 1**). Eighty-two participants were in all three datasets.

**Table 1.** Characteristics of Sheffield participants by dataset. Continuous variables are median (interquartile range); categorical variables are n/N (%); denominators below the column total reflect missing data. Plasma and CSF MND columns include one pre-symptomatic *C9orf72* carrier coded as MND at assay, who was not analysed. Deceased, recorded date of death (database snapshot 8 December 2025). Disease stage, onset to sampling divided by onset to death. ALS, amyotrophic lateral sclerosis; ALSFRS-R, revised ALS Functional Rating Scale; CSF, cerebrospinal fluid; FVC, forced vital capacity; MND, motor neurone disease; NA, not applicable or not recorded; PBP, progressive bulbar palsy; PLS, primary lateral sclerosis; PMA, progressive muscular atrophy.

| Characteristic | Proteome, MND (n = 118) | Proteome, ALS (n = 94) | ALS stage set (n = 75) | Proteome, controls (n = 24) | Plasma, MND (n = 95) | Plasma, controls (n = 39) | CSF, MND (n = 76) | CSF, controls (n = 34) |
|---|---|---|---|---|---|---|---|---|
| MND variant, ALS/PMA/PLS/PBP | 94/14/9/1 | 94/0/0/0 | 75/0/0/0 | NA | 75/11/7/0 | NA | 60/10/4/0 | NA |
| Age at consent, years | 62.6 (56.4 to 71.4) | 62.5 (56.4 to 71.4) | 65.2 (58.0 to 72.3) | 63.0 (54.8 to 69.2) | 62.7 (56.5 to 70.2) | 58.0 (50.5 to 66.5) | 62.1 (55.0 to 70.1) | 57.0 (48.5 to 65.8) |
| Male | 63/118 (53.4) | 48/94 (51.1) | 37/75 (49.3) | 5/24 (20.8) | 49/93 (52.7) | 14/39 (35.9) | 39/74 (52.7) | 13/34 (38.2) |
| Site of onset, limb/bulbar/other | 97/16/5 | 76/13/5 | 59/12/4 | NA | 73/16/4 | NA | 59/12/3 | NA |
| Bulbar onset | 16/118 (13.6) | 13/94 (13.8) | 12/75 (16.0) | NA | 16/93 (17.2) | NA | 12/74 (16.2) | NA |
| Familial | 12/118 (10.2) | 12/94 (12.8) | 11/75 (14.7) | NA | 7/93 (7.5) | NA | 5/74 (6.8) | NA |
| *C9orf72* expansion | 8/118 (6.8) | 8/94 (8.5) | 8/75 (10.7) | NA | 9/95 (9.5) | NA | 6/76 (7.9) | NA |
| Onset to sampling, months | 17.0 (10 to 26) | 14.5 (9 to 24) | 13.0 (8.5 to 21.5) | NA | 17.0 (10 to 25) | NA | 15.0 (9.2 to 25) | NA |
| ALSFRS-R at sampling (n) | 38 (32 to 42) (105) | 38 (30.5 to 41) (83) | 36 (30 to 40) (67) | NA | 39 (32 to 42) (79) | NA | 40 (34 to 42.2) (68) | NA |
| FVC, % predicted (n) | 86.5 (70.8 to 101) (100) | 89 (69 to 102) (77) | 85 (62 to 99) (61) | NA | 89 (70.5 to 103) (82) | NA | 90 (74.5 to 103.5) (67) | NA |
| Progression rate, ALSFRS-R points per month | 0.6 (0.3 to 1.1) | 0.7 (0.4 to 1.2) | 0.9 (0.5 to 1.5) | NA | 0.5 (0.3 to 1.0) | NA | 0.5 (0.3 to 1.0) | NA |
| Deceased | 85/118 (72.0) | 75/94 (79.8) | 75/75 (100) | NA | 69/93 (74.2) | NA | 53/74 (71.6) | NA |
| Onset to death, months (deceased) | 34 (24 to 53) | 33 (21.5 to 46) | 33 (21.5 to 46) | NA | 35 (25 to 55) | NA | 34 (24 to 49) | NA |
| Disease stage (deceased) | 0.4 (0.3 to 0.6) | 0.5 (0.3 to 0.6) | 0.5 (0.3 to 0.6) | NA | 0.4 (0.3 to 0.7) | NA | 0.4 (0.3 to 0.7) | NA |

The 94 participants with ALS in the proteomic analysis set had a median age at sampling of 62.5 years (interquartile range (IQR) 56.4 to 71.4); 48 (51.1%) were male, 13 (13.8%) had bulbar onset, 12 (12.8%) had a family history of ALS and 8 (8.5%) carried a *C9orf72* repeat expansion. Median onset-to-sampling time was 14.5 months, median ALSFRS-R at sampling was 38 (IQR 30.5 to 41) and median progression rate was 0.7 ALSFRS-R points per month. By 8 December 2025, 75 (79.8%) had died, at a median of 33 months after onset; these 75 deceased patients form the Sheffield disease-stage analysis set (median stage at sampling 0.46). Of the 94, 64 also had plasma and 50 CSF metabolomes.

Findings were replicated in three independent cohorts with survival or longitudinal data: Tel Aviv (serum mass spectrometry; 349 patients with ALS, 290 deceased),^9^ Oxford (serum Olink Explore HT; 381 patients, 260 deceased)^10^ and AnswerALS (plasma SomaScan; 129 deceased patients with ALS and 431 samples)^18^ (**Supplementary Table 2**; **Methods**).

### Circulating proteins associated with ALS survival time

In 94 participants with ALS from Sheffield (75 deaths; median time from onset to death or censoring 38.1 months, IQR 25.8 to 67.3), the serum concentration of 199 proteins was nominally associated with survival time (107 with higher and 92 with lower hazard, **Figure 2A; Supplementary Table 3**). In this dataset only NfL passed a multiple testing correction (hazard ratio (HR) per standard deviation (SD) 2.67, 95% CI 1.78 to 3.99, P = 1.9 × 10⁻⁶, FDR = 0.010, **Figure 2A**), with no departure from proportional hazards (P = 0.44).

**Figure 2.**
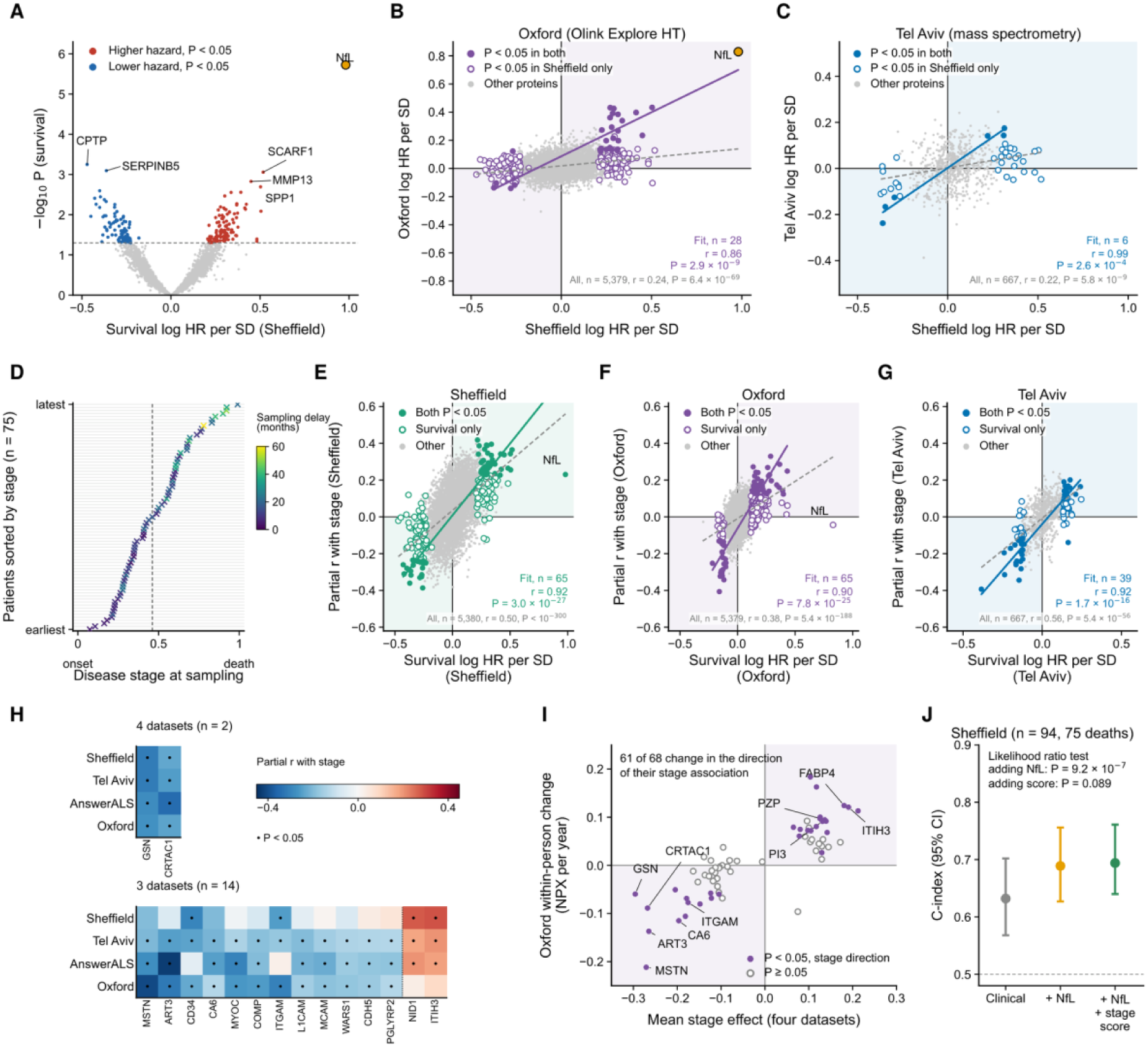
Survival-associated proteins replicate across cohorts and are enriched for associations with disease stage. **(A)** Survival associations of 5,401 proteins in 94 Sheffield participants with ALS (75 deaths; Cox models of time from onset with delayed entry at sampling, adjusted for age at sampling and sex). Red and blue, higher and lower hazard at P < 0.05; gold, NfL (the only protein at FDR < 0.05); dashed line, P = 0.05. (**B, C**) Survival log HR per SD in Sheffield against Oxford (B; Olink Explore HT; 5,379 shared proteins; 381 participants, 260 deaths) and Tel Aviv (**C**; mass spectrometry; 667 shared proteins; 343 participants, 288 deaths). Coloured, Sheffield survival-associated proteins (P < 0.05): filled, also P < 0.05 in the replication cohort; open, P < 0.05 in Sheffield only. Grey, other proteins; gold, NfL. Shading, quadrants of concordant direction. Coloured line, linear fit to the filled proteins (P < 0.05 in both cohorts); grey line, linear fit to all proteins; n, r and P shown for each. (**D**) Disease stage at sampling in 75 deceased Sheffield patients with ALS (one row per patient, sorted by stage; 0, onset; 1, death). Crosses are coloured by sampling delay (months from symptom onset to sampling); dashed line, median stage (0.46). (**E–G**) Survival log HR per SD against partial correlation with disease stage (adjusted for age and sex) within each dataset: Sheffield (E; 94 participants for survival, 75 deceased for stage), Oxford (F; 381 and 260) and Tel Aviv (G; 343 and 290). Filled, P < 0.05 for both survival and stage; open, survival P < 0.05 only; grey, other proteins. Coloured line, linear fit to the filled proteins; grey line, linear fit to all proteins; n, r and P shown for each. Shading, quadrants in which higher hazard corresponds to higher levels at later stage. (**H**) Partial correlation with disease stage of the 16 stage-associated proteins with P < 0.05 in all four datasets (top; GSN and CRTAC1) or in three (bottom; 14 proteins), in Sheffield, Tel Aviv, AnswerALS (129 deceased participants with ALS, 431 samples) and Oxford (rows); dot, P < 0.05. The 52 proteins with P < 0.05 in two datasets are shown in Supplementary Figure 3. (**I**) Within-person change of the 68 stage-associated proteins in 222 Oxford participants with repeated samples (668 samples; fixed-effects slope of NPX on months since onset, cluster-robust standard errors, expressed per year) against their mean stage effect across the four datasets. Filled, within-person slope P < 0.05 in the direction of the stage association; open, P ≥ 0.05. Shading, concordant quadrants. (**J**) Optimism-corrected Harrell’s C-index (95% CI; 200 bootstrap resamples) for survival in Sheffield (94 participants, 75 deaths; time from onset with delayed entry) with clinical covariates (age at sampling, sex and bulbar onset; grey), with NfL added (orange) and with NfL and the 68-protein stage score added (green). The stage score is the mean of z-scored protein levels, each multiplied by the sign of its stage association. P values are from likelihood ratio tests for each addition; dashed line, 0.5. ALS, amyotrophic lateral sclerosis; FDR, false discovery rate; HR, hazard ratio; NfL, neurofilament light chain; NPX, Normalized Protein eXpression; SD, standard deviation.

Proteins nominally associated with survival time in Sheffield are also significantly associated with survival time in Oxford and in Tel Aviv (**Figure 2B, C**). All 199 Sheffield survival-associated proteins were measured in the Oxford cohort: 131 (65.8%) had the same direction of effect (binomial P = 9.4 × 10⁻⁶; **Figure 2B**), and 28 had P < 0.05 in Oxford (10.0 expected; binomial P = 8.7 × 10⁻⁷), 27 of them in the same direction. These proteins were more likely to be associated with survival in Oxford than the remaining 5,180 proteins (14.1% against 3.6%; odds ratio 4.37, 95% CI 2.86 to 6.69; Fisher’s exact P = 3.3 × 10⁻⁹). Across the proteome, log hazard ratios correlated between Sheffield and Oxford (r = 0.24, 95% CI 0.21 to 0.26, P = 6.4 × 10⁻⁶⁹; 5,379 proteins), and more strongly among the 199 Sheffield survival-associated proteins (r = 0.48, 95% CI 0.37 to 0.58, P = 7.2 × 10⁻¹³; **Figure 2B**). Thirty-seven of the 199 proteins were measured in the Tel Aviv cohort; reduced consistency might be expected because Sheffield and Oxford proteomes were assayed on the same platform (Olink Explore HT), whereas Tel Aviv used mass spectrometry. Twenty-nine (78%) had the same direction of effect in Tel Aviv (binomial P = 7.5 × 10⁻⁴), and 6 had P < 0.05, all concordant (1.9 expected; binomial P = 0.0095), although this rate did not differ significantly from that of other proteins (16.2% against 11.9%; odds ratio 1.43, P = 0.44). Log hazard ratios correlated between Sheffield and Tel Aviv across 667 shared proteins (r = 0.22, 95% CI 0.15 to 0.29, P = 5.8 × 10⁻⁹) and among the 37 Sheffield survival-associated proteins (r = 0.67, 95% CI 0.44 to 0.82, P = 5.9 × 10⁻⁶; **Figure 2C**).

The 199 Sheffield survival-associated circulating proteins are independently supported as disease-specific by studies of ALS cases and controls. Of the 199 survival-associated proteins, 24 were among the 270 measured proteins that differ between ALS and controls in Ran et al.^8^ (137 proteins) or Chia et al.^7^ (203 proteins at FDR < 0.05). Nine were in both sets. This was more than the 9.9 expected by chance across the 5,401 proteins measured (odds ratio 2.76, one-sided Fisher’s exact P = 4.5 × 10⁻⁵). In 23 of the 24, the direction of the difference between ALS and controls matched the direction of the association with hazard (binomial P = 3.0 × 10⁻⁶) (**Supplementary Figure 1A**).

### ALS survival-associated proteins carry a disease-stage signal

To derive disease stage we normalised the total disease duration for each patient to be 0 at symptom onset and 1 at death (**Figure 1A, Methods**). Median stage at sampling was 0.46 in Sheffield (75 deceased patients with ALS) but the complete spectrum of disease stage is represented (**Figure 2D**); median stage at sampling is 0.45 in Tel Aviv (290), 0.51 in Oxford (260) and 0.38 in AnswerALS (129 patients, 431 draws) (**Supplementary Figure 2**). Disease stage is not simply sampling delay (**Figure 2D**).

Serum protein concentrations nominally associated with survival in Sheffield were also associated with disease stage in Sheffield, Oxford and Tel Aviv (**Figure 2E–G; Supplementary Table 4**). In Sheffield, 184 of the 199 survival-associated proteins (92.5%) had the same direction of association with stage as with hazard (higher hazard with higher levels at later stage; binomial P = 3.7 × 10⁻³⁸), and 65 had stage P < 0.05 (10.0 expected; binomial P = 7.5 × 10⁻³⁵), all in the same direction (**Figure 2E**). These proteins were more likely to be stage-associated than the remaining 5,181 proteins (32.7% against 8.7%; odds ratio 5.08, 95% CI 3.72 to 6.93; Fisher’s exact P = 2.8 × 10⁻²⁰). Because disease stage incorporates survival time, we tested the same 199 proteins against stage in independent cohorts. In Oxford, 131 (65.8%) had the same direction of association with stage (binomial P = 9.4 × 10⁻⁶), and 30 had stage P < 0.05 (10.0 expected; binomial P = 7.8 × 10⁻⁸), 25 of them in the same direction (**Figure 2F**); they were more likely to be stage-associated than the remaining 5,180 proteins (15.1% against 5.8%; odds ratio 2.86, 95% CI 1.91 to 4.29; Fisher’s exact P = 6.0 × 10⁻⁶). Of the 37 proteins measured in Tel Aviv, 30 (81%) had the same direction of association with stage (binomial P = 1.9 × 10⁻⁴) and 11 had stage P < 0.05 (1.9 expected; binomial P = 1.2 × 10⁻⁶), 10 of them in the same direction (**Figure 2G**), although this proportion did not differ significantly from that of other proteins (29.7% against 21.4%; odds ratio 1.55, 95% CI 0.75 to 3.22, P = 0.23).

### Proteome-wide analysis of ALS disease stage derives a set of 68 reproducible stage proteins

We have established that circulating prognostic proteins are enriched with stage-associated proteins. Next, we aimed to establish a set of reproducible stage-associated circulating proteins for further characterisation.

Among the 542 circulating proteins where concentration was measured in all four proteomic datasets, stage associations agreed between cohorts: partial correlations with stage correlated in all six pairwise comparisons (r = 0.19 to 0.45, all P < 1 × 10⁻⁵), and the direction of association agreed in 91% of pairwise comparisons (**Methods**). Sixty-eight proteins were nominally associated with stage in at least two datasets (31 higher and 37 lower at later stage, **Figure 2H**; **Supplementary Figure 3; Supplementary Table 5**) which is more than would be expected by chance (binomial P = 1.3 × 10⁻⁸; permutation P < 0.001, 2,000 permutations). Weighting for censoring did not alter observed stage correlations (r = 0.992 with unweighted values in Sheffield and 0.980 in Tel Aviv). NfL was associated with stage in Sheffield at borderline statistical significance (r = 0.23, P = 0.046) but not in AnswerALS (0.07, P = 0.29) or Oxford (−0.04, P = 0.48); we conclude that NfL is not a stage-associated protein.

The 68 stage-associated circulating proteins are independently supported as disease-specific by studies of ALS cases and controls. Of the 68 stage-associated proteins, 18 were in the Ran et al.^8^ or Chia et al.^7^ ALS-versus-control sets. This is more than the 6.9 expected among the 542 proteins measured in all four ALS proteomics datasets (odds ratio 4.25, one-sided Fisher’s exact P = 2.6 × 10⁻⁵), and seven were in both sets. All 18 differed between ALS and controls in the same direction as with later stage (binomial P = 7.6 × 10⁻⁶; Ran et al. 8 of 8, P = 0.0078; Chia et al. 17 of 17, P = 1.5 × 10⁻⁵). CA3 and ITIH3 were higher in ALS, and the other 16, including CRTAC1, COMP, MYOC, MSTN and GSN, were lower (**Supplementary Figure 1B**).

### Longitudinal changes in stage protein concentrations

As an internal validity check for the 68 proteins associated with ALS-disease stage across multiple datasets, we determined the direction and magnitude of change in circulating concentration in a subset of ALS patients where longitudinal measurements were available.

To test whether stage associations observed between patients reflect change within patients, we estimated the within-person slope of each protein over time in 222 Oxford participants with repeated samples (668 samples; fixed-effects regression of NPX on months since onset with cluster-robust standard errors). Sixty-one of the 68 stage-associated proteins (90%) changed over time in the direction of their stage association (binomial P = 7.4 × 10⁻¹²), with a median slope of +0.072 NPX per year for proteins higher and −0.029 NPX per year for proteins lower at later stage (**Figure 2I; Supplementary Table 6**). The magnitude of within-person change tracked the strength of the cross-sectional stage association (r = 0.84 between within-person slope and mean stage effect across the four datasets, P = 6.0 × 10⁻¹⁹).

### ALS survival prediction is improved by incorporating stage protein measurements

We have identified a set of 68 circulating proteins associated with ALS-disease stage across multiple datasets. We hypothesised that incorporating these markers into a prediction model for ALS survival time would add performance compared to a model using only NfL which is not a disease-stage marker.

For each participant, a stage score was calculated from the 68 stage-associated proteins (**Methods**), so that higher scores indicate a more advanced proteomic stage. In Sheffield (94 participants with ALS, 75 deaths), nested Cox proportional hazards models of time from onset to death were fitted using age at sampling (per 10 years), sex, site of disease onset, and with/without NfL and stage score. Discrimination was assessed with Harrell’s concordance index (C-index), corrected for optimism by 200 bootstrap resamples, with 95% CIs from the same resamples; the contribution of each added term was tested by likelihood ratio test. The C-index was 0.632 (95% CI 0.568 to 0.702) with clinical covariates and rose to 0.689 (0.627 to 0.756) with NfL added (likelihood ratio χ² = 24.1, P = 9.2 × 10⁻⁷). Adding the stage score raised it to 0.694 but this change was not statistically significant (0.640 to 0.761; likelihood ratio χ² = 2.9, P = 0.089; stage score HR per SD 1.25, 95% CI 0.97 to 1.60, **Figure 2J**).

### Gelsolin is a reproducible biomarker of ALS disease stage and is prognostic when combined with NfL

We have introduced the idea that ALS disease stage is a distinct prognostic entity with a different biological basis from the rate of neuroaxonal injury exemplified by circulating concentration of NfL. Among the 68 proteins which make up our disease stage panel (**Figure 2H**) we hypothesised that circulating gelsolin (GSN) may be a disease stage biomarker because it exhibited a consistent and significant stage association in all four ALS proteomics datasets. Moreover, gelsolin has been identified separately as a prognostic biomarker for ALS.^9^

Among the 542 proteins measured in all four cohorts, gelsolin had the strongest association with disease stage: the largest mean and the largest minimum absolute partial correlation across cohorts. It fell with stage in Sheffield (partial r = −0.32, P = 0.0057; n = 75), Tel Aviv (r = −0.31, P = 4.6 × 10⁻⁸; n = 290), AnswerALS (r = −0.28, P = 5.9 × 10⁻⁵; n = 129) and the Oxford cohort (r = −0.27, P = 9.6 × 10⁻⁶; n = 260), so gelsolin alone explained 7.3% to 10.0% of the variance in stage (**Supplementary Figure 4A**). In contrast NfL was nominally associated with stage in Sheffield (r = 0.23, P = 0.046) but not in AnswerALS (r = 0.07, P = 0.29) or Oxford (r = −0.04, P = 0.48); Tel Aviv did not measure NfL (**Supplementary Figure 4A**). In 222 Oxford participants with repeated samples (668 samples), gelsolin fell by 0.060 NPX per year within individuals (within-individual fixed-effects regression, P = 0.0055; **Figure 2I**), so its association with stage holds over time within the same person as well as across participants.

Gelsolin and NfL each tracked one dimension of disease. In deceased participants with both measures, NfL explained 26% of the variance in ALSFRS-R decline in Oxford (n = 221) and 33% in Sheffield (n = 67), against < 1% and 1% of the variance in stage. In contrast, gelsolin explained 16% and 8% of the variance in stage, against 2% and < 1% of that in decline (partial Spearman ρ², adjusted for age, sex, centre and the other dimension; **Figure 1A**; **Figure 3A**). Decline and stage were almost unrelated (Spearman ρ = −0.19 in Oxford and −0.02 in Sheffield). Mean NfL rose across quartiles of decline (z = −0.65 to +0.83) but not of stage, whereas gelsolin fell across quartiles of stage (+0.38 to −0.38) and less across quartiles of decline (+0.19 to −0.15) (**Figure 3B**).

**Figure 3.**
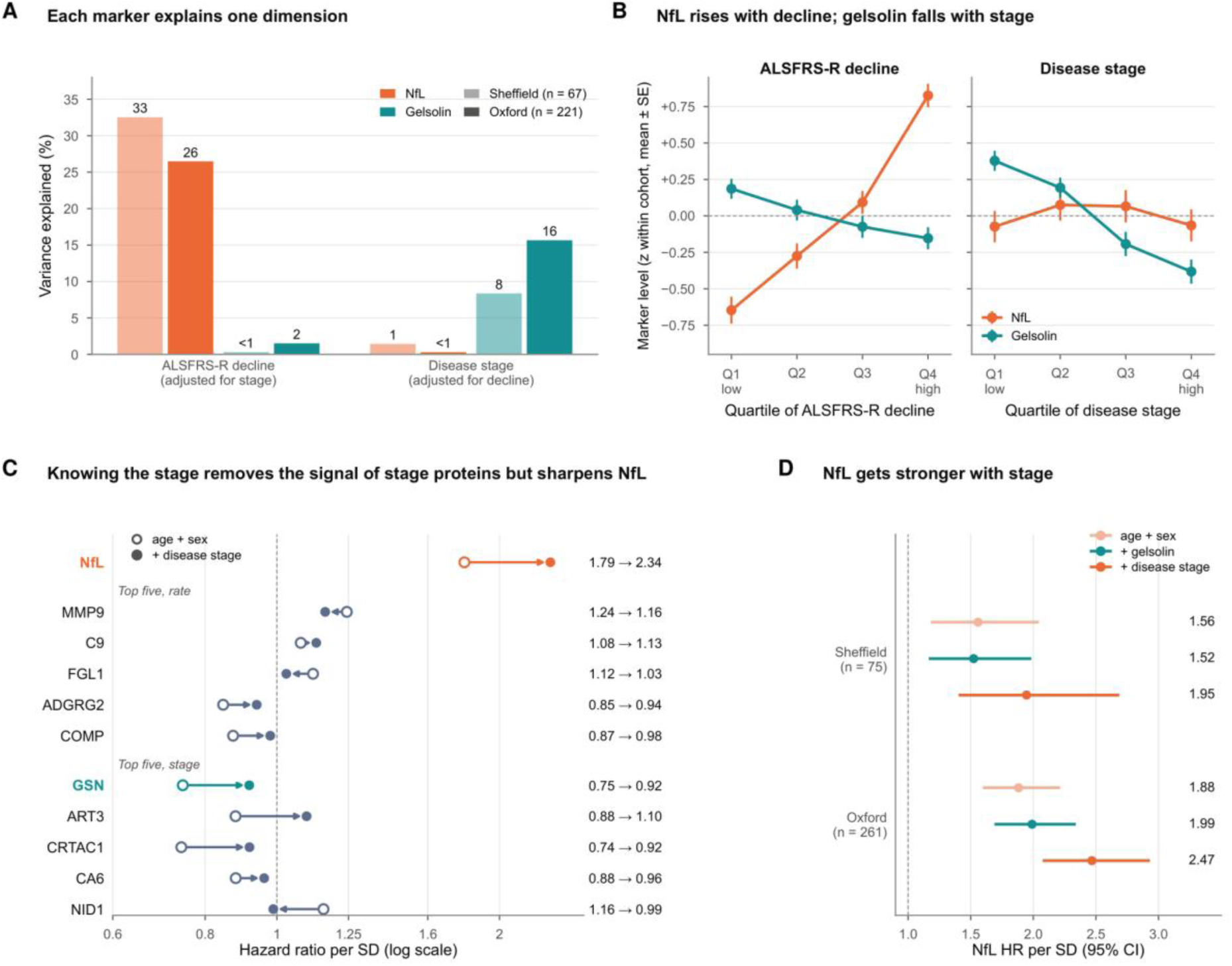
Gelsolin marks disease stage, NfL the rate of decline. (**A**) Variance in ALSFRS-R decline (adjusted for stage) and in disease stage (adjusted for decline) explained by NfL (orange) and gelsolin (teal) in deceased participants with both measures, in Sheffield (light; n = 67) and Oxford (full; n = 221). Partial Spearman ρ², adjusted for age, sex and centre. (**B**) Marker level (z within cohort, mean ± SE) by quartile of ALSFRS-R decline (left) and of disease stage (right). NfL, Sheffield and Oxford; gelsolin, Sheffield, Oxford and Tel Aviv. (**C**) Cox HR per SD for survival from sampling in deceased participants, adjusted for age and sex (open circles) and with disease stage added (filled circles), for NfL and for the five most rate-associated and five most stage-associated of the 68 stage proteins. Proteins, Sheffield and Tel Aviv; NfL, Sheffield and Oxford; inverse-variance pooled. ART3 crosses 1. The increase for NfL exceeds that of 98% of stage-unrelated proteins in Sheffield, so it is not explained by non-collapsibility alone. (**D**) NfL HR per SD (95% CI) in deceased participants in Sheffield (n = 75) and Oxford (n = 261), adjusted for age and sex (light orange), with gelsolin added (teal) and with disease stage added (orange). ALSFRS-R decline, (48 − ALSFRS-R)/months since onset. Dashed lines, HR = 1. ALSFRS-R, revised ALS Functional Rating Scale; HR, hazard ratio; NfL, neurofilament light chain; SD, standard deviation.

NfL carries prognostic information independent of disease position, whereas the prognostic signal in gelsolin and other stage proteins is largely accounted for by stage (**Figure 3C,D**). Adding stage slightly strengthened the association of NfL with survival time: absolute log HR +2.8% in Sheffield (HR per SD 2.17, 95% CI 1.60 to 2.94, without stage; 2.21, 1.62 to 3.03, with stage, **Supplementary Table 7**) and +2.7% in Oxford. It weakened the association of gelsolin with survival time by 16.1% in Sheffield, 22.6% in Tel Aviv (0.88, 0.78 to 0.99, P = 0.03, without stage; 0.91, 0.80 to 1.02, P = 0.11, with stage, **Supplementary Table 7**) and 49.6% in the Oxford cohort (**Supplementary Table 7**).

To this point we have used a definition of disease stage that requires survival time. To validate gelsolin as a biomarker we tested a different measure of disease stage: the D50 model which calculates the time taken to lose half of functional capacity as measured using longitudinal ALSFRS-R scores (**Supplementary Figure 4B**).^6^ Here we define “aggressiveness” (−log D50, speed of decline) which is analogous to the rate of neuroaxonal injury, and disease phase (rD50, the fraction of the course to the functional midpoint elapsed at sampling) which is analogous to disease stage. In 170 Oxford participants with at least two ALSFRS-R scores (median D50 41.2 months, IQR 24.9 to 59.9) gelsolin tracked phase (standardised β −0.41, 95% CI −0.54 to −0.28, P = 8.4 × 10⁻⁹) but not aggressiveness (−0.08, −0.21 to 0.05, P = 0.25) (**Supplementary Figure 4C**). In contrast NfL tracked aggressiveness (0.56, 0.43 to 0.68, P = 1.1 × 10⁻¹⁴) but not phase (0.13, −0.003 to 0.26, P = 0.056) (**Supplementary Figure 4C**). Mean gelsolin fell across tertiles of phase (z = 0.49, 0.04 and −0.53) and mean NfL rose across tertiles of aggressiveness (−0.60, −0.04 and 0.64) (**Supplementary Figure 4D**). Across all 4,050 Olink assays, gelsolin ranked second for association with phase but NfL had the strongest association with aggressiveness (**Supplementary Figure 4E; Supplementary Table 8**). Gelsolin explained 19% of the variance in phase (partial R², adjusted for aggressiveness, age, sex and study, **Supplementary Figure 4C**).

Together, gelsolin and NfL improved survival prediction. In Sheffield and Oxford pooled (475 participants, 336 deaths), the cross-validated C-index for survival from sampling was 0.60 (95% CI 0.57 to 0.64) with age and sex, 0.63 (0.60 to 0.66) with gelsolin, 0.73 (0.70 to 0.76) with NfL and 0.76 (0.73 to 0.78) with both (**Figure 1A**). Gelsolin added 0.027 (95% CI 0.014 to 0.042) to NfL (likelihood ratio P = 7 × 10⁻¹²; joint HR per SD, NfL 2.31 and gelsolin 0.66), with the same pattern for pairs within each dataset (0.72 to 0.75).

### ALS stage-associated proteins change with ageing and disease progression is associated with an increase in proteomic age gap

We have identified a set of 68 proteins where the circulating concentration is associated with ALS-disease stage. Next, we sought to understand the biological basis of these proteins and therefore of ALS disease progression (**Figure 1B**).

We hypothesised that circulating stage proteins might be associated with clinical variables known to determine prognosis in ALS: age at symptom onset, sex and site of onset. In 94 participants with ALS from Sheffield, each of the 68 stage-associated proteins was regressed on age at onset, sex and bulbar onset jointly (**Figure 4A**). Age at onset was associated with 23 proteins at P < 0.05 (3.4 expected; binomial P = 1.1 × 10⁻¹³); 21 of the 23 were higher with older age (binomial P = 6.6 × 10⁻⁵ for direction). Sex was associated with 16 proteins (binomial P = 1.9 × 10⁻⁷; 10 higher in men), and bulbar onset with 11 (binomial P = 5.3 × 10⁻⁴; 8 higher with bulbar onset) (**Supplementary Table 9**). The strong and largely unidirectional association with age led us to test whether part of the stage signal reflects accelerated proteomic ageing.

**Figure 4.**
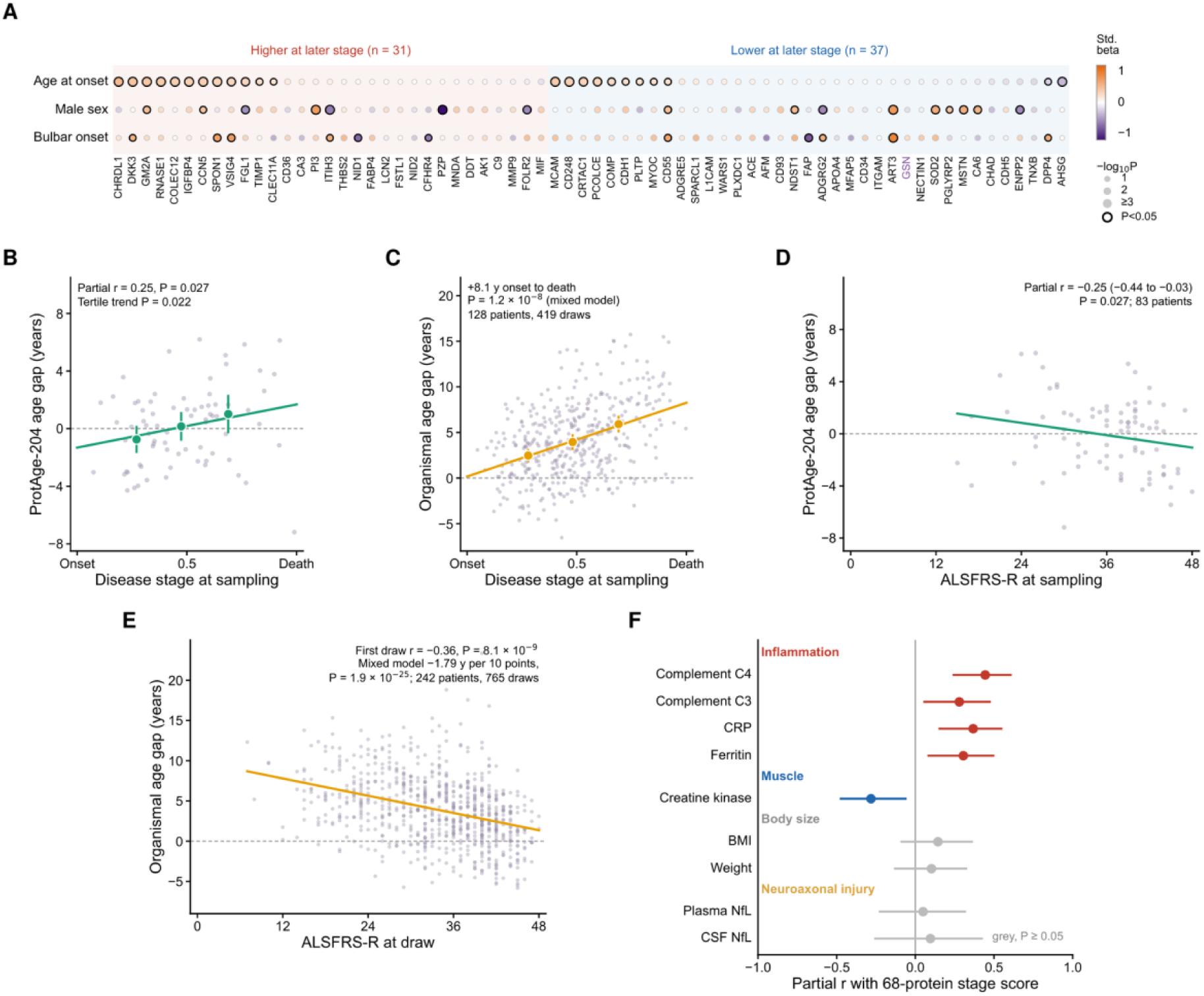
Clinical determinants of the stage proteins, and proteomic age gap against disease stage and function. **(A)** Clinical determinants of the 68 stage proteins in Sheffield patients with ALS (n = 94). Each dot is the standardised coefficient (colour; purple, negative; orange, positive) from a multivariable linear regression of the protein on age at onset, sex and site of onset. Dot size scales with −log₁₀ P, and a black outline marks P < 0.05. Proteins are grouped by whether they are higher (n = 31) or lower (n = 37) at later disease stage and ordered by the age-at-onset effect; gelsolin (GSN) is highlighted. **(B)** Proteomic age gap (ProtAge minus chronological age, years; residual on age at sampling) against disease stage in deceased Sheffield patients with ALS (n = 75). Points show patients, large points stage-tertile means (95% confidence interval) and the line a linear fit. **(C)** Organismal proteomic age gap (Oh et al., 2023; SomaScan, years, adjusted for plate) against disease stage in AnswerALS (128 deceased patients with ALS, 419 draws). Points show draws, large points stage-tertile means (95% confidence interval) and the line a linear fit; P from a mixed model with a random intercept per patient. **(D)** Proteomic age gap (ProtAge-204, years) against ALSFRS-R at sampling in Sheffield patients with ALS (n = 83). Points show patients and the line a linear fit; partial Spearman correlation adjusted for age and sex. **(E)** Organismal proteomic age gap (Oh et al., 2023; SomaScan, years, adjusted for plate) against ALSFRS-R at the same visit in AnswerALS patients with ALS (242 patients, 765 draws). Points show draws and the line the fixed effect of a mixed model with a random intercept per patient, adjusted for age and sex; the partial Spearman correlation uses the first draw per patient (n = 239). **(F)** Partial Pearson correlation between the 68-protein stage score (signed mean z-score) and laboratory and clinical measures recorded at the sampling visit in Sheffield patients with ALS, adjusted for age at sampling and sex. Points show r and lines show 95% confidence intervals (Fisher z transformation). Coloured points have P < 0.05 and grey points P ≥ 0.05; n = 34–75 per measure. Two weights were corrected for decimal-point errors and CRP below detection was set to 0.2 mg/L.

To externally validate our hypothesis that disease-stage proteins are associated with age, we used UK Biobank plasma proteomic data which were generated with the Olink Explore 3072 platform (2,923 proteins) in plasma samples from 54,219 participants in the UK Biobank Pharma Proteomics Project.^19^ To generate a set of age-associated proteins the circulating concentration of each protein was correlated with age at recruitment (Spearman, FDR < 0.05, |ρ| ≥ 0.10, **Methods**).

Of the 68 stage-associated proteins, 67 were measured in UK Biobank (WARS1 was not). Thirty-four of the 67 were associated with age in UK Biobank (Spearman, FDR < 0.05 and |ρ| ≥ 0.10), against 27.5 expected among the 510 proteins measured in all four ALS proteomic datasets and UK Biobank (one-sided Fisher’s exact test, P = 0.054, **Supplementary Table 10**). Across these 510 proteins, the association with age in UK Biobank is correlated with the calculated ALS stage association (Pearson r = 0.19, P = 1.9 × 10⁻⁵), and more strongly among the 67 stage-associated proteins (r = 0.32, P = 0.0073). Agreement depended on the direction of the stage association (**Supplementary Table 10**). All 17 shared proteins that were higher at later stage were also higher with older age, whereas only 2 of the 17 shared proteins that were lower at later stage were lower with older age. Proteins that rise with disease stage therefore resemble population proteomic ageing, whereas proteins that fall with stage change in the opposite direction to normal ageing.

Finally, proteomic age estimated with the 204-protein ProtAge clock, retrained in UK Biobank with the published code and protein list,^17^ rose nominally with disease stage in Sheffield: the proteomic age gap increased from −0.75 years in the earliest to +1.01 years in the latest stage tertile (partial Spearman correlation adjusted for age and sex, r = 0.25, P = 0.027, BH-adjusted P = 0.055 across four clocks; test for trend P = 0.022) (**Figure 4B; Supplementary Table 10**). The association was similar with NfL removed from the clock (r = 0.21, P = 0.069). SImilarly a clock trained on SomaScan (Oh et al., 2023) showed a rise in age gap with stage in AnswerALS (+8.1 years from onset to death, P = 1.2 × 10⁻⁸; **Figure 4C; Supplementary Figure 5C**). The age gap was also larger in patients with lower ALSFRS-R at sampling, in Sheffield (partial Spearman r = −0.25, 95% CI −0.44 to −0.03, P = 0.027; n = 83; **Figure 4D**) and in AnswerALS with the SomaScan-trained clock (first draw r = −0.36, P = 8.1 × 10⁻⁹; 1.8 years per 10 ALSFRS-R points lost across repeated draws, P = 1.9 × 10⁻²⁵; **Figure 4E**).

### ALS stage-associated proteins implicate systemic inflammation and muscle atrophy

We have identified a set of 68 proteins where the circulating concentration is associated with ALS-disease stage and with an acceleration of biological aging. To further understand the stage signature, we correlated the 68-protein stage score with laboratory and anthropometric measures recorded at the sampling visit in Sheffield.

As before we used calculated stage score as the signed mean z-score of the 68 proteins. We used partial Pearson correlation adjusted for age at sampling and sex (**Figure 4F; Supplementary Table 11**). A higher stage score was associated with higher complement C4 (partial r = 0.44, 95% CI 0.24–0.61, P = 9.4 × 10⁻⁵; n = 74), C-reactive protein (r = 0.37, 95% CI 0.15–0.55, P = 0.0016; n = 73), ferritin (r = 0.30, 95% CI 0.08–0.50, P = 0.0097; n = 73) and complement C3 (r = 0.28, 95% CI 0.05–0.48, P = 0.018; n = 74). It was also associated with lower creatine kinase (r = −0.28, 95% CI −0.48 to −0.06, P = 0.016; n = 75). Complement C4 and C-reactive protein remained significant after Bonferroni correction for nine tests (threshold P < 0.0056). The score was not associated with body mass index (r = 0.14, 95% CI −0.09 to 0.37, P = 0.24; n = 72) or weight (r = 0.10, 95% CI −0.14 to 0.33, P = 0.40; n = 72). Nor was it associated with plasma neurofilament light (r = 0.05, P = 0.74; n = 52) or cerebrospinal fluid neurofilament light (r = 0.10, P = 0.60; n = 34). Together these results link later disease stage to a systemic acute-phase inflammatory response and reduced muscle mass, independent of weight loss and of neuroaxonal injury.

### ALS disease stage associated plasma and CSF metabolites reflect the same underlying biology as circulating proteins

Survival associations of these plasma and CSF metabolites have been reported previously.^11^ Here we tested association with disease stage, using the same model as for proteins, in deceased Sheffield patients with ALS (**Methods**). In plasma (n = 60), 149 of 1,271 named metabolites were associated with stage at P < 0.05 (64 expected, binomial P = 2.4 × 10⁻²¹). In CSF (n = 46), 58 of 436 were associated (22 expected, P = 2.0 × 10⁻¹¹) (**Figure 5A; Supplementary Table 13**). Sixteen metabolites were associated with stage in both fluids, and 15 of these changed in the same direction. These included three collagen-derived metabolites (prolylhydroxyproline, hydroxyproline and galactosylhydroxylysine), five sphingomyelins and acetylspermidine, all of which rose with stage (**Figure 5B**).

**Figure 5.**
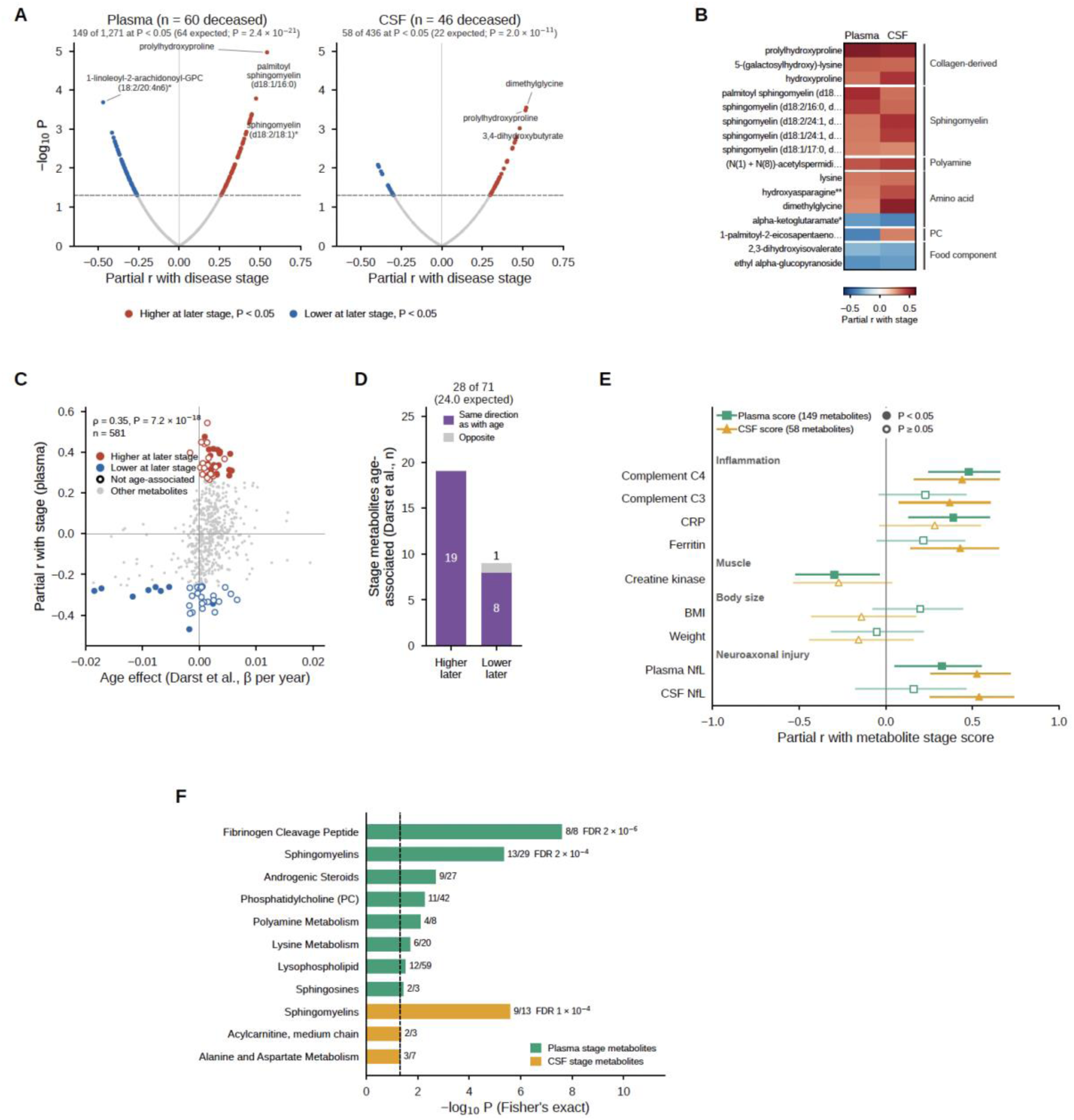
Circulating metabolites associated with ALS stage. **(A)** Partial correlation of each named metabolite with disease stage, adjusted for age at sampling and sex, in deceased Sheffield patients with ALS: plasma (n = 60) and CSF (n = 46). Red marks metabolites higher and blue marks metabolites lower at later stage (P < 0.05). The number at P < 0.05 is compared with the number expected (binomial test). **(B)** Stage partial r in plasma and CSF for the 16 metabolites associated with stage (P < 0.05) in both fluids, grouped by class. **(C)** Age effect from a population study (Darst et al., β per year) plotted against stage partial r in plasma for 581 matched metabolites. Stage-associated metabolites are coloured by direction. Filled points are age-associated (Bonferroni P < 0.05) and open points are not. **(D)** Among the 28 plasma stage-associated metabolites that were also age-associated, the number whose stage effect is in the same direction as their age effect (purple) or the opposite direction (grey), split by direction of change with stage. **(E)** Partial correlation (95% CI), adjusted for age at sampling and sex, of the plasma stage score (149 metabolites; squares) and the CSF stage score (58 metabolites; triangles) with laboratory and clinical measures recorded at sampling. Filled markers indicate P < 0.05. **(F)** Over-representation of Metabolon sub-pathways among stage-associated metabolites in plasma (green) and CSF (orange). One-sided Fisher’s exact test against all named metabolites in each fluid. Shown are sub-pathways with nominal P < 0.05. Labels give the number of stage-associated metabolites out of the sub-pathway size, and FDR where it is below 0.05. The dashed line marks P = 0.05.

Like circulating proteins, stage-associated plasma metabolites tended to change in the direction of ageing. Across 581 plasma metabolites matched to a population study of age including 2,344 samples from 1,212 dementia-free participants,^20^ stage associations correlated with age effects (Spearman ρ = 0.35, P = 7.2 × 10⁻¹⁸, **Figure 5C**). Of 71 matched stage-associated metabolites, 28 were age-associated (24.0 expected; odds ratio 1.33, P = 0.17). In 27 of these 28, the direction of change with stage matched the direction of change with age (binomial P = 2.2 × 10⁻⁷, **Figure 5D**).

As for circulating proteins, we built a metabolite stage score for each fluid (the signed mean z-score of its stage-associated metabolites) and related it to laboratory measures recorded at sampling. The plasma score correlated with complement C4 (partial r = 0.48, P = 1.3 × 10⁻⁴) and CRP (r = 0.33, P = 0.015), and inversely with creatine kinase (r = −0.30, P = 0.025) (**Figure 5E; Supplementary Table 11**). The CSF score correlated with complement C4 (r = 0.44, P = 0.0023), complement C3 (r = 0.31, P = 0.037) and ferritin (r = 0.44, P = 0.0029) (**Figure 5E**). Neither score was associated with body mass index or weight. Unlike the protein score, both metabolite scores correlated with plasma NfL (plasma r = 0.33, P = 0.017; CSF r = 0.53, P = 3.3 × 10⁻⁴), and the CSF score also correlated with CSF NfL (r = 0.54, P = 5.7 × 10⁻⁴) suggesting that disease stage defined by metabolites is not entirely independent of the rate of neuroaxonal injury. We tested stage-associated metabolites for over-representation in Metabolon sub-pathways^21^. Consistent with an inflammatory signal, plasma stage-associated metabolites were over-represented among fibrinogen cleavage peptides (8 of 8; Fisher’s exact test, FDR = 2 × 10⁻⁶) and sphingomyelins (13 of 29; FDR = 2 × 10⁻⁴). Sphingomyelins were also over-represented in CSF (9 of 13; FDR = 1 × 10⁻⁴) (**Figure 5F; Supplementary Table 14**).

## Discussion

We set out to separate two components of the circulating prognostic signal in ALS: how fast the disease progresses, and how far it has progressed at the time of sampling. We defined disease stage as the fraction of the onset-to-death interval elapsed at sampling. Serum proteins associated with survival in Sheffield were strongly enriched for stage associations including in external cohorts. Sixty-eight proteins tracked stage across four cohorts which we defined as a panel of stage-associated circulating proteins. They changed over time within individuals in the direction of their stage association, and published ALS-versus-control studies independently identify them as disease-related.^7,8^ The circulating protein stage signature reflected an acute-phase inflammatory response, loss of muscle, and a pattern of change that resembles, and in part accelerates, proteomic ageing. Plasma and CSF metabolites associated with stage reflected the same inflammatory and ageing biology. Throughout, NfL, the established prognostic biomarker in ALS,^3,4^ behaved differently from the stage-associated proteins.

### Disease rate and disease position are distinct prognostic dimensions

A biomarker can be prognostic because it reflects the rate of neurodegeneration, or because it reflects position within the disease course. Patients sampled later in their course die sooner, so a marker of position is prognostic even if it is unrelated to disease aggressiveness. Disease stage includes survival time in its denominator, so part of the correlation between survival and stage effects is built in. Two observations argue that the stage signal we have detected is not an artefact of this construction i.e. that prognostic proteins and metabolites are *not* coincidentally associated with disease stage. First, stage-associated proteins changed over time within individuals in the direction of their stage association, a comparison that does not use survival time. Second, the D50 phase measure^6^ is derived from functional decline without reference to survival, and it reproduced the stage associations.

NfL tracked the rate of functional decline estimated with the D50 model^6^ but not phase which describes the proportion of functional decline occurred at a given time point and is analogous to disease stage. The 68 stage-associated proteins we have identified showed the opposite pattern: stage attenuated their survival associations, and in the D50 model they tracked phase but not aggressiveness. Our observations with respect to NfL agree with other studies using the D50 model: CSF NfL tracked aggressiveness but not phase.^22^ Serum NfL tracked aggressiveness while troponin T tracked phase, both in two independent German cohorts^23^ and in a Swedish population-based cohort.^24^

Normalising time to the individual disease course has precedent. Milestones of King’s clinical staging occur at consistent fractions of the onset-to-death interval,^5,25^ and motor cortical dysfunction has been staged in the same way.^26^ To our knowledge, this is the first application of such a measure to the circulating proteome and metabolome in ALS.

### Gelsolin is a reproducible biomarker of disease stage

Of the 542 proteins measured in all four cohorts, gelsolin (GSN) had the strongest and most consistent association with stage, and it was one of only two proteins associated with stage in every cohort. On its own, gelsolin explained 7–10% of the between-patient variance in stage, across three proteomic platforms and in both serum and plasma. It explained 19% of the variance in D50 phase, a measure that does not depend on survival time. Gelsolin declined within individuals sampled repeatedly, and it was not associated with age at onset, sex or site of onset. These effect sizes are comparable to those that established NfL as a marker of disease rate. In the same Oxford participants and models, NfL explained 30% of the variance in the rate of functional decline (-log D50). In an independent serum study, NfL correlated with progression rate at r = 0.34, or about 11% of the variance.^27^ Gelsolin therefore explains a share of the variance in disease position similar to the share of variance in progression rate explained by NfL. NfL is now used for prognostic stratification and as a pharmacodynamic marker in trials.^4,28^

Other studies also implicate gelsolin in ALS. A combined analysis of 25 proteomic studies found it consistently dysregulated across ALS biofluids.^29^ An earlier mass spectrometry study found it lower in ALS plasma than in controls,^30^ as did both of the published ALS-versus-control proteomes used here.^7,8^ It was also among the prioritised prognostic proteins identified in Tel Aviv.^9^ Plasma gelsolin is an abundant component of the extracellular actin-scavenging system,^31^ so it may be measurable with simpler single-analyte assays than proteomic platforms. That would allow it to be tested as a stand-alone stage marker alongside NfL. Together, the two markers could indicate from a single blood sample how fast a patient is progressing (NfL) and how far they have progressed (gelsolin). Both quantities matter for trial stratification and for interpreting change on treatment.

### Biology of the stage signature: inflammation, muscle and ageing

A higher stage-protein score was associated with higher complement C4, C3, C-reactive protein and ferritin, and with lower creatine kinase. It was not associated with body mass index, weight or NfL. Later disease therefore carries a systemic acute-phase inflammatory signature and a fall in creatine kinase consistent with loss of muscle mass, rather than a stronger neuroaxonal injury signal. These laboratory measures have been evaluated as prognostic markers in ALS,^28^ but our data suggest that part of their prognostic signal may reflect the stage at which they were sampled. Proteins lower at later stage included myostatin, myocilin, COMP, CRTAC1 and GSN. This is consistent with the involvement of skeletal muscle reported years before symptom onset.^7^ The metabolome pointed to the same biology. Stage-associated plasma metabolites were over-represented among fibrinogen cleavage peptides, which are released when thrombin cleaves the acute-phase protein fibrinogen. They were also over-represented among sphingomyelins, the most consistent class across plasma and CSF, which longitudinal metabolomics in ALS has also implicated.^12^ Collagen-derived metabolites (prolylhydroxyproline, hydroxyproline and galactosylhydroxylysine) rose with stage in both fluids, consistent with turnover of connective tissue and muscle. Unlike the protein score, the metabolite stage scores correlated with NfL, particularly in CSF. Disease stage defined by metabolites is therefore not fully independent of the rate of neuroaxonal injury.

The stage-associated proteins also overlapped with ageing. All 17 proteins that rose with stage and were age-associated in UK Biobank also rose with age,^19^ and the proteomic age gap estimated with a published clock^17^ increased nominally with disease stage. Plasma metabolites associated with stage, whether they rose or fell, changed in the direction of population ageing.^20^ Ageing is the strongest risk factor for ALS. Our findings suggest that disease progression is accompanied by a systemic shift toward an older molecular profile, although these data cannot show whether this shift is a cause or a consequence of advancing disease.

### Implications for prognosis and biomarker discovery

In Sheffield, a 68-protein stage score added little to survival prediction beyond NfL. The more important implication is for interpretation: proteins identified as prognostic, or as differing between patients and controls, may in part reflect when patients were sampled. All 18 stage-associated proteins found in published ALS-versus-control sets differed in the direction of later stage.^7,8^ Accounting for stage in study design and analysis would separate markers of disease rate, which are candidate pharmacodynamic markers, from markers of disease position, which are candidate stratification variables.

### Limitations

Disease stage can be defined only in deceased patients and includes survival time. Because stage includes survival time, adjustment for stage partly conditions on the outcome, so attenuation should not be interpreted as mediation. Oxford results were returned as aggregates, and the D50 analysis used a simplified model in 170 participants. The 68-protein signature was defined without correction for multiple testing. Metabolite analyses were confined to Sheffield, with no independent metabolomic cohort. Laboratory measures were recorded at a single visit and were available only for subsets of participants. Fasting status and sample-handling variables were not recorded.

## Conclusion

### Disease stage offers an explanation for many of the prognostic proteins previously linked to ALS

Circulating prognostic signals in ALS combine markers of how fast the disease progresses with markers of how far it has progressed. GSN is a reproducible single-protein marker of disease stage and complements NfL as a marker of rate. Its association with stage is comparable in size to the association of NfL with rate, which makes it a candidate for translation as a stratification test.

## Methods

### Participants and ethics

All data come from the Sheffield arm of the AMBRoSIA study (A Multicentre Biomarker Resource Strategy In ALS),^28^ (hereafter Sheffield). The study was approved by the London South East Research Ethics Committee (16/LO/2136); the serum assayed here is stored under the Motor System Genomics approval (12/YH/0330). All participants gave written informed consent. People with motor neurone disease (MND; ALS, progressive muscular atrophy (PMA), primary lateral sclerosis (PLS) or progressive bulbar palsy (PBP)) were recruited between 6 June 2017 and 6 December 2023. Controls were volunteers without neurodegenerative disease. Analyses were restricted to participants with ALS (94 in the proteomic analysis set; **Table 1**).

### Clinical and laboratory data

The pseudonymised clinical database records sex, age at consent, MND variant, familial status, site and date of onset, C9orf72 and SOD1 status, revised ALS Functional Rating Scale (ALSFRS-R) total,^32^ forced vital capacity (FVC, % predicted) and date of death. ALSFRS-R item scores and gastrostomy and ventilation dates were not recorded, so King’s and MiToS stages^5,33^ were not derived. Survival analyses used the export of 8 December 2025, with living participants censored on that date.

Blood (serum separator, EDTA and lithium heparin tubes) and CSF (lumbar puncture; polypropylene tubes) were collected at the baseline (consent) visit, centrifuged at 3,500 rpm for 10 min at 4 °C within 1 h and stored in vapour-phase nitrogen.^28^ Fasting status, time of day, time to processing, clotting time, storage temperature and freeze-thaw history were not recorded. Serum was stored for a median of 7.0 years (range 4.4 to 8.5) before assay. The following were recorded at the sampling visit: serum concentration of complement C3 and C4, C-reactive protein (CRP), ferritin, creatine kinase (CK), weight and body mass index (BMI), plasma and CSF neurofilament light chain (NfL). Complement values recorded in mg/dL were converted to g/L. Two weights recorded as 6305 and 5805 kg were corrected to 63.05 and 58.05 kg (decimal-point errors) and BMI recalculated; BMI values below 12 or above 70 kg/m² were otherwise treated as missing. CRP below the detection limit was set to 0.2 mg/L, half the lowest recorded value. CK, CRP, ferritin and both NfL measures were log-transformed.

### Proteomic profiling

Serum was profiled with Olink Explore HT^34,35^ by the Oxford Multiomics Technology Platforms, Wellcome Centre for Human Genetics (project P250108; Illumina NovaSeq 6000; NPX Map 2.0.0). Two plates each held 86 sample and 10 control wells (**Supplementary Table 1**). Fifteen of 5,440 assays (5,416 proteins and 24 internal controls) failed vendor quality control, leaving 5,401. We used intensity-normalised Normalized Protein eXpression (NPX, log2 scale), and no detection filter was applied.

NPX was z-scored per assay across all 172 sample wells, including pooled-replicate and control wells, so hazard ratios (HR) are per 172-well standard deviation (SD) (within-ALS SD 0.35 to 1.33). The proteomic analysis set holds 118 MND participants (94 ALS, 14 PMA, 9 PLS, 1 PBP), excluding one participant with probable MND recorded as off study.

### Metabolomic profiling

EDTA plasma (n = 134) and CSF (n = 110) were profiled by Metabolon (Morrisville, NC, USA; project SHEF-0101-24DSML+) on the Global Discovery (HD4) platform.^21,36^ The platform uses four ultra-high-performance liquid chromatography tandem mass spectrometry methods and more than 5,400 authentic standards, and reports peak area. Each fluid ran on four plates, with patients and controls on every plate, and as one batch per method; data were median-scaled.

Released matrices are median-scaled and minimum-imputed (plasma) or extracted-volume-normalised (CSF), on the linear scale. No sample or feature was removed; zero-variance features and features with > 40% of values at the imputed floor are flagged. One participant coded as MND at shipment is a pre-symptomatic C9orf72 carrier, included in assayed counts (95 MND in plasma, 76 in CSF) but not analysed. Analyses used named metabolites (excluding unnamed “X-” features), log2-transformed and annotated with Metabolon super-and sub-pathways.^21^

### Replication cohorts and published datasets

Proteomes were measured in serum in Sheffield, Tel Aviv and the Oxford cohort, and in plasma in AnswerALS, UK Biobank and the published ALS-versus-control studies (**Supplementary Table 2**).

#### Tel Aviv

Mass spectrometry proteomes and clinical data from 349 people with ALS (290 deceased).^9^ The stage analysis used 1,219 proteins and the survival analysis 1,218 proteins with ≥ 20 values in 343 participants (288 deaths).

#### AnswerALS

SomaScan v4.1 (7,596 aptamers; 7,274 against human proteins)^37,38^ for 943 samples from 358 participants at up to five time points, with onset, death and draw dates and ALSFRS-R.^18^

#### Oxford cohort

Olink Explore HT proteomes (5,416 proteins) and clinical data from people with ALS.^10^ Oxford investigators ran our analysis scripts on individual-level data and returned aggregate results only, with counts < 5 suppressed. Analyses used the first sample of 381 participants (253 male; 260 deceased); 106 of them had also been recruited at the Oxford site of the same multicentre study.

#### UK Biobank

Olink Explore 3072 plasma proteomes (2,923 proteins) from 54,219 participants in the UK Biobank Pharma Proteomics Project.^19,39^

#### Published ALS-versus-control protein sets

From Ran et al.,^8^ 137 Olink Explore HT proteins differing between manifest ALS (n = 35) and controls (n = 59) at false discovery rate (FDR) < 0.05 (105 higher, 32 lower in ALS). From Chia et al.,^7^ all 203 proteins at FDR < 0.05 among 2,868 assays (Olink Explore 3072).

#### Population metabolite ageing

Age effects for plasma metabolites from Darst et al.,^20^ 2,344 samples from 1,212 dementia-free participants.

### Disease stage

Disease stage was months from onset to sampling divided by months from onset to death (0, onset; 1, death), in deceased patients only. Samples dated at or after death were assigned a stage of 0.99 in every cohort; in Sheffield this affected one participant. Analyses used 75 deceased patients with ALS in Sheffield, 290 in Tel Aviv, 260 in the Oxford cohort and 129 in AnswerALS (431 draws; four participants with non-ALS MND excluded).

### Protein survival analyses

In 94 participants with ALS (75 deaths), each protein was entered into a Cox proportional hazards model of time from onset to death with delayed entry at sampling (left truncation)^40^ to exclude immortal time,^41,42^ adjusted for age at sampling and sex (R survival package).^43^ FDR was controlled across 5,401 assays. Proportional hazards were assessed with scaled Schoenfeld residuals. Sensitivity analyses used age at onset, no delayed entry, or entry from recorded dates (94 participants, 75 deaths; NfL HR 2.58, FDR = 0.026).

The same model was fitted in Tel Aviv (343 participants, 288 deaths; 1,218 proteins scaled per SD) and the Oxford cohort (381 participants, 260 deaths; 5,415 proteins; adjusted for age at onset). Sign agreement between cohorts was tested by two-sided binomial test, and log HRs were correlated (Pearson). Enrichment of Sheffield survival-associated proteins (P < 0.05) among proteins with P < 0.05 in each replication cohort was tested by Fisher’s exact test.

### Stage association analyses

Stage associations were partial Pearson correlations adjusted for sex and age (at sampling in Sheffield and Oxford; at onset in Tel Aviv). They were estimated in Sheffield (5,401 proteins), Tel Aviv (proteins with ≥ 20 values) and the Oxford cohort (5,415 proteins).

In AnswerALS, which has repeated samples per participant, log10 relative fluorescence units were regressed on stage, age, sex and bulbar onset across all draws. The stage coefficient was estimated after residualising protein level and stage on the covariates (Frisch–Waugh–Lovell), with standard errors clustered by participant and t tests on n − 5 degrees of freedom, where n is the number of participants. Where several aptamers targeted the same protein, the aptamer with the lowest P value was retained.

Among the 542 proteins measured in all four datasets, the stage signature comprised proteins with P < 0.05 and the same direction in ≥ 2 datasets, without correction for multiple testing. The number expected by chance was obtained from the binomial distribution given the number of stage-associated proteins in each dataset, and from 2,000 permutations of protein labels within each dataset. Sheffield and Tel Aviv correlations were recomputed with inverse probability of censoring weights.

### Within-person longitudinal change in the Oxford cohort

In 222 Oxford participants with repeated samples (668 samples), the within-person (fixed-effects) slope of NPX on months since onset was estimated with cluster-robust standard errors. Agreement with the direction of the stage association was tested by two-sided binomial test. Within-person slopes were correlated (Pearson) with mean stage effects across the 68 stage-associated proteins.

### Overlap with published ALS-versus-control proteins

Overlaps with the combined Ran et al. and Chia et al. ALS-versus-control sets were tested by one-sided Fisher’s exact test against all proteins measured in the datasets defining each set (survival, 5,401 proteins; stage, the 542 proteins measured in all four cohorts). Direction concordance with published ALS-versus-control effects was tested by two-sided binomial test.

### Stage score and survival prediction

For each participant, a stage score was calculated as the mean of the z-scored levels of the 68 stage-associated proteins, each multiplied by the sign of its stage association. Higher scores therefore indicate a more advanced proteomic stage.

In Sheffield (94 participants, 75 deaths), nested Cox models of time from onset with delayed entry were fitted in three steps: age at sampling (per 10 years), sex and bulbar onset; NfL added; the stage score added. Discrimination was Harrell’s C-index,^44^ corrected for optimism by 200 bootstrap resamples, with 95% CIs from the same resamples. Nested models were compared by likelihood ratio test.

### Clinical determinants of the stage-associated proteins

In 94 participants with ALS, each stage-associated protein (z-scored) was regressed on z-scored age at onset, male sex and bulbar onset. For each covariate, the number of proteins with P < 0.05 was compared with the 5% expected by one-sided binomial test.

### Population ageing and proteomic age

In UK Biobank, each protein was correlated with age at recruitment (Spearman), and age-associated proteins were defined as FDR < 0.05 and |ρ| ≥ 0.10. Overlap of the stage-associated proteins with UK Biobank age-associated proteins was tested by one-sided hypergeometric test against the 510 proteins measured in all four ALS datasets and UK Biobank. UK Biobank age correlations were correlated (Pearson) with mean stage effects, across these 510 proteins and within the stage-associated proteins. Agreement of direction was tabulated separately for proteins higher and lower at later stage.

Age correlations in Sheffield, Tel Aviv and AnswerALS are reported in **Supplementary Table 10**. They were computed with Spearman correlation in all MND participants in Sheffield, 349 participants in Tel Aviv, and 308 participants at the first time point in AnswerALS (best aptamer per gene).

Proteomic age was estimated with the 204-protein ProtAge clock,^17^ retrained because the fitted model was not released: we used the authors’ code and protein list in 44,573 UK Biobank participants without MND (LightGBM, proteins only, 70/30 split, hyperparameters tuned with Optuna; held-out r = 0.93, mean absolute error 2.31 years). Sheffield Olink Explore HT values were z-scored per protein within the 149 Sheffield samples and rescaled to the UK Biobank mean and SD (196 of 204 proteins measured). The proteomic age gap was the residual of predicted age on age at sampling, fitted in the 94 patients with ALS. In the 75 deceased patients with ALS, the gap was related to stage by partial Spearman correlation adjusted for age and sex, and summarised by stage tertile with a test for linear trend. The analysis was repeated with NfL removed from the clock, with ProtAge20 and with a clock trained on the 65 ageing-signature proteins; P values were adjusted by the Benjamini-Hochberg method across these four clocks.

### Laboratory correlates and Gene Ontology over-representation

In deceased Sheffield participants with ALS, the stage score was correlated with each laboratory and body-size measure by partial Pearson correlation adjusted for age at sampling and sex. A Bonferroni threshold for nine tests (P < 0.0056) was applied.

Over-representation of GO biological process terms was tested by one-sided Fisher’s exact test, with Benjamini-Hochberg FDR within each protein set. The analysis used: Library: Gene Ontology Biological Process 2023 as distributed by Enrichr;^45^ Terms: restricted to inflammation, immunity or muscle terms by keyword matching of term names (keyword lists in **Supplementary Table 12**); Term size: 5 to 300 proteins within the 542-protein background; Protein sets: all 68 stage-associated proteins, the 31 higher at later stage and the 37 lower at later stage; Testable terms: those containing at least two proteins from the set being tested.

### Gelsolin and NfL

Gelsolin (GSN) was selected because, among the 542 proteins measured in all four cohorts, it had the largest mean and the largest minimum absolute partial correlation with stage. The variance in stage explained by a single protein was the squared partial correlation.

Survival associations of NfL and GSN were compared without and with stage as an additional covariate. These Cox models of time from onset were adjusted for age at sampling and sex (Oxford, age at onset), in Sheffield (n = 75), Tel Aviv (n = 288) and the Oxford cohort (n = 260; HR per NPX unit). Attenuation was the percentage change in absolute log HR on adding stage; the comparison was repeated for all proteins (**Supplementary Table 7**).

ALSFRS-R decline at sampling was (48 − ALSFRS-R)/months from onset to sampling. In deceased participants with both measures (Sheffield, n = 67; Oxford first sample, n = 221), the variance in decline and in stage explained by NfL or gelsolin was the squared partial Spearman correlation (linear regression on ranks), adjusted for age, sex, Oxford centre and the other dimension. For quartile plots, markers were z-scored within cohort and pooled (NfL, Sheffield and Oxford; gelsolin, also Tel Aviv).

Stage-adjusted survival associations used Cox models of survival from sampling in deceased participants, adjusted for age and sex (Oxford, stratified by centre), without and with stage, or with gelsolin for NfL. Protein HRs were pooled across Sheffield and Tel Aviv, and NfL across Sheffield and Oxford, by inverse-variance weighting. The five most rate- and stage-associated of the 68 proteins were ranked by partial correlation with decline or stage combined across Sheffield, Tel Aviv and AnswerALS (FDR < 0.05 for 5 proteins with decline and 44 with stage). To allow for non-collapsibility of the HR, the change in absolute log HR of NfL on adding stage in Sheffield was compared with that of proteins weakly correlated with stage (|r| < 0.1).

Pooled survival prediction used Sheffield participants with ALS and the first Oxford sample per participant (475 participants, 336 deaths). Cox models of survival from sampling were stratified by dataset (Sheffield and five Oxford centres), with age, NfL and gelsolin z-scored within cohort. Models contained age and sex, with gelsolin, NfL or both added. Harrell’s C-index was estimated by 5 repeats of 10-fold cross-validation (folds stratified by dataset and event), with 95% CIs and the gain from gelsolin from 500 bootstrap resamples. Nested models were compared by likelihood ratio test on the full data.

### Disease aggressiveness and phase in the Oxford cohort

Oxford investigators shared NfL and gelsolin NPX, ALSFRS-R, covariates, and onset, sampling and event intervals for 823 samples from 388 participants with ALS (none were Sheffield participants). Sampling times after the event (15 samples) or exceeding the onset-to-event interval (22) were set to 0.99 of that interval.

In 170 participants with ≥ 2 ALSFRS-R scores, a simplified D50 model was fitted by nonlinear least squares: a sigmoid from 48 at onset to 24 at D50, the months from onset to loss of half the score. Aggressiveness was z-scored −log D50, and phase (rD50) was z-scored months from onset to first sample divided by D50. First-sample NfL and gelsolin (z-scored) were regressed jointly on aggressiveness and phase, adjusted for age at sampling, sex and study. Partial R² for each term was derived from its t statistic as t²/(t² + residual degrees of freedom, 161). In 261 deceased participants, each protein was regressed on z-scored stage with the same covariates.

Sensitivity analyses used: participants with ≥ 3 scores (n = 85); aggressiveness as z-scored 1/D50; a linear mixed model of all 550 samples (random intercept; adjusted for age and sex; phase at each sample).

### Metabolite stage analyses

In deceased Sheffield participants with ALS (plasma n = 60; CSF n = 46), each named metabolite with ≥ 20 non-missing values was correlated with disease stage, adjusted for age at sampling and sex (partial Pearson correlation; plasma 1,271 metabolites, CSF 436). The number with P < 0.05 was compared with the 5% expected by one-sided binomial test. Overlap of stage-associated metabolites between fluids was tested by one-sided Fisher’s exact test among metabolites measured in both, and stage correlations were compared between fluids (Pearson). Survival associations of these metabolites have been reported separately^11^ and were not re-analysed.

Plasma stage correlations were compared (Spearman) with published age effects (581 metabolites matched by name).^20^ Age-associated metabolites were those passing a Bonferroni threshold across the 581 matched metabolites. Their enrichment among stage-associated metabolites (P < 0.05) was tested by Fisher’s exact test, and agreement in direction between stage and age by two-sided binomial test.

For each biofluid, a metabolite stage score was calculated as the mean of the z-scored stage-associated metabolites, each multiplied by the sign of its stage association (plasma 149, CSF 58). The scores were correlated with laboratory measures as for the protein score. Over-representation of stage-associated metabolites in Metabolon sub-pathways with ≥ 3 measured metabolites was tested by one-sided Fisher’s exact test, with Benjamini-Hochberg FDR within each fluid (plasma 96, CSF 46 sub-pathways).

### Statistical analysis

Tests were two-sided at P < 0.05 unless stated; FDR was controlled by the Benjamini-Hochberg procedure. Python: numpy, scipy, statsmodels, lifelines and matplotlib. R 4.6.0: survival 3.8.6 (Efron ties), ggplot2 4.0.3, patchwork 1.3.2, dplyr 1.2.1, tidyr 1.3.2, readr 2.2.0, readxl 1.5.0, forcats 1.0.1, ggrepel 0.9.8, scales 1.4.0 and ragg 1.5.2.

## Author contributions

P.J.S, J.CK, A.S and M.W conceptualised the work with assistance from E.H and H.K. A.S and M.W wrote and executed the code for the project with assistance from C.H, H.K, and Y.C. A.D organised sample sequencing. M.B. collected and collated patient metadata. P.J.S oversaw the Sheffield arm of the study. J.C.K, A.S, F.M., I.M., Y.C., D.G.L., A.G.T., and M.W wrote the manuscript; all authors assisted with manuscript drafting. J.CK, A.T, E.H, S.Z and P.J.S supervised the work and edited the manuscript

## Acknowledgements

We are very grateful to the patients with ALS and control subjects who generously donated biosamples and to UKB participants. This research was conducted using the UK Biobank Resource under Application Number 68812. Figures were created using BioRender.com. Olink analysis of the Oxford cohort was supported by GSK through the Oxford-GSK Institute of Molecular and Computational Medicine. We acknowledge the support of the NIHR Oxford Biomedical Research Centre.

## Funding

This work was supported by My Name’5 Doddie Foundation (DOD/14/43 to PJS) and NIHR (NF-SI-0617–10077 to PJS). The authors would like to acknowledge the Deanship of Graduate Studies and Scientific Research, Taif University, for funding this work. JCK is supported by the MND Association (894–791) and TargetALS. We acknowledge the support of the NIHR Sheffield Biomedical Research Centre (NIHR 203321) and the NIHR Sheffield Clinical Research Facility.

## Competing Interests

Kari Wong was an employee of Metabolon at the time of the work described.

## Data availability

Anonymised metabolic and proteomic profiling data from the AMBRoSIA project can be provided upon request by a qualified researcher.

## Supplementary Material

**Supplementary Figure 1.**
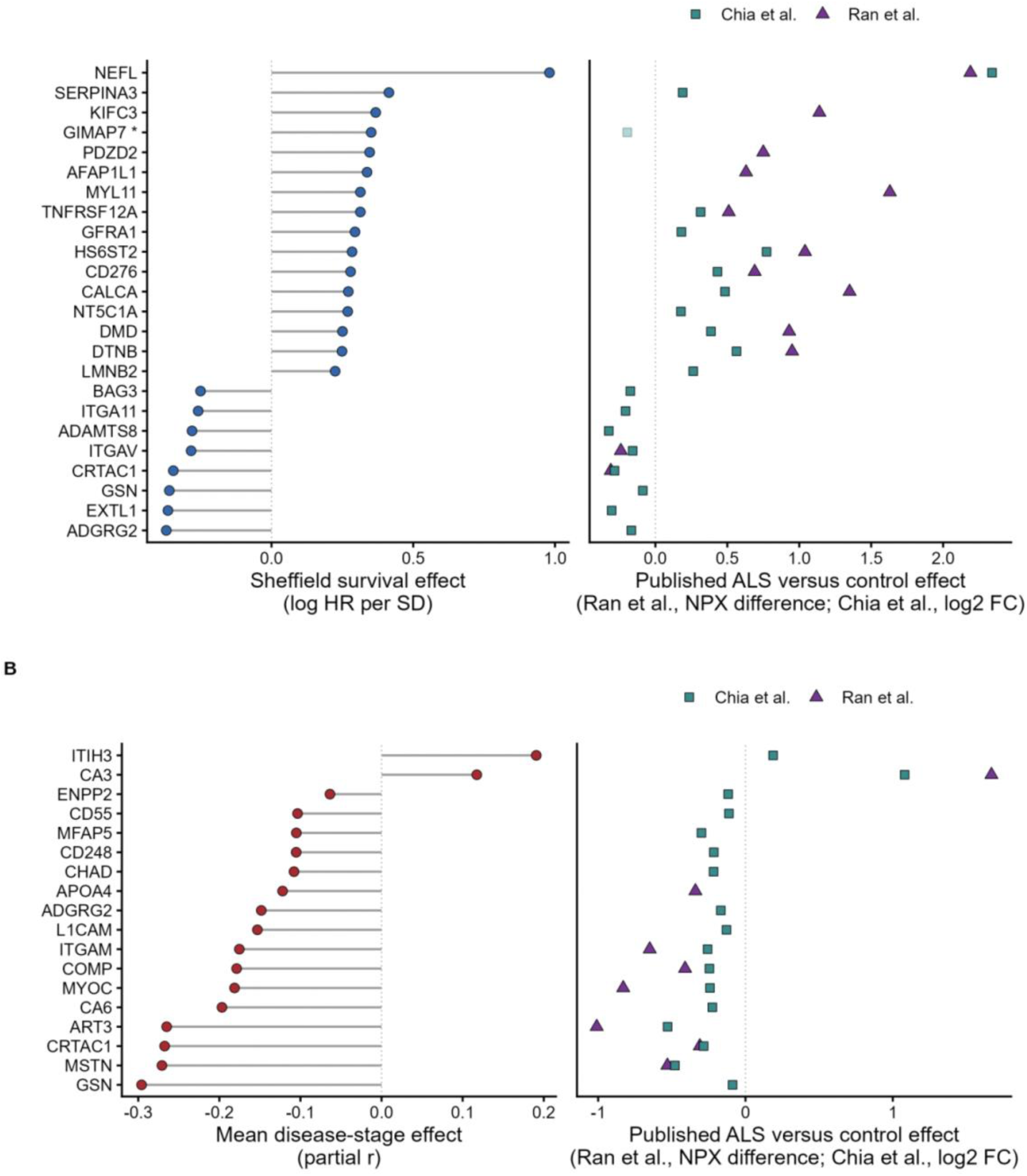
Survival- and stage-associated proteins in published ALS-versus-control proteomes. (**A**) The 24 Sheffield survival-associated proteins (P < 0.05) that also differ between ALS and controls in Ran et al.^8^ (137 proteins; Olink Explore HT) or Chia et al.^7^ (203 proteins at FDR < 0.05; Olink Explore 3072). The left plot shows the Sheffield survival effect (log HR per SD), from a Cox model of time from onset with delayed entry at sampling, adjusted for age at sampling and sex. The right plot shows the published ALS-versus-control effect: NPX difference for Ran et al. (triangles) and log2 fold change for Chia et al. (squares). Proteins in both published sets are shown with both symbols. An asterisk and a faded symbol mark a protein whose published difference is in the opposite direction to its survival association. (**B**) The 18 of the 68 stage-associated proteins that are in the Ran et al. or Chia et al. ALS-versus-control sets. The left plot shows the mean disease-stage effect: partial r, adjusted for age and sex, averaged across Sheffield, Tel Aviv, AnswerALS and the Oxford cohort. The right plot shows the published ALS-versus-control effect, as in A. Dotted lines mark no effect. ALS, amyotrophic lateral sclerosis; FC, fold change; HR, hazard ratio; NPX, normalised protein expression; SD, standard deviation.

**Supplementary Figure 2.**
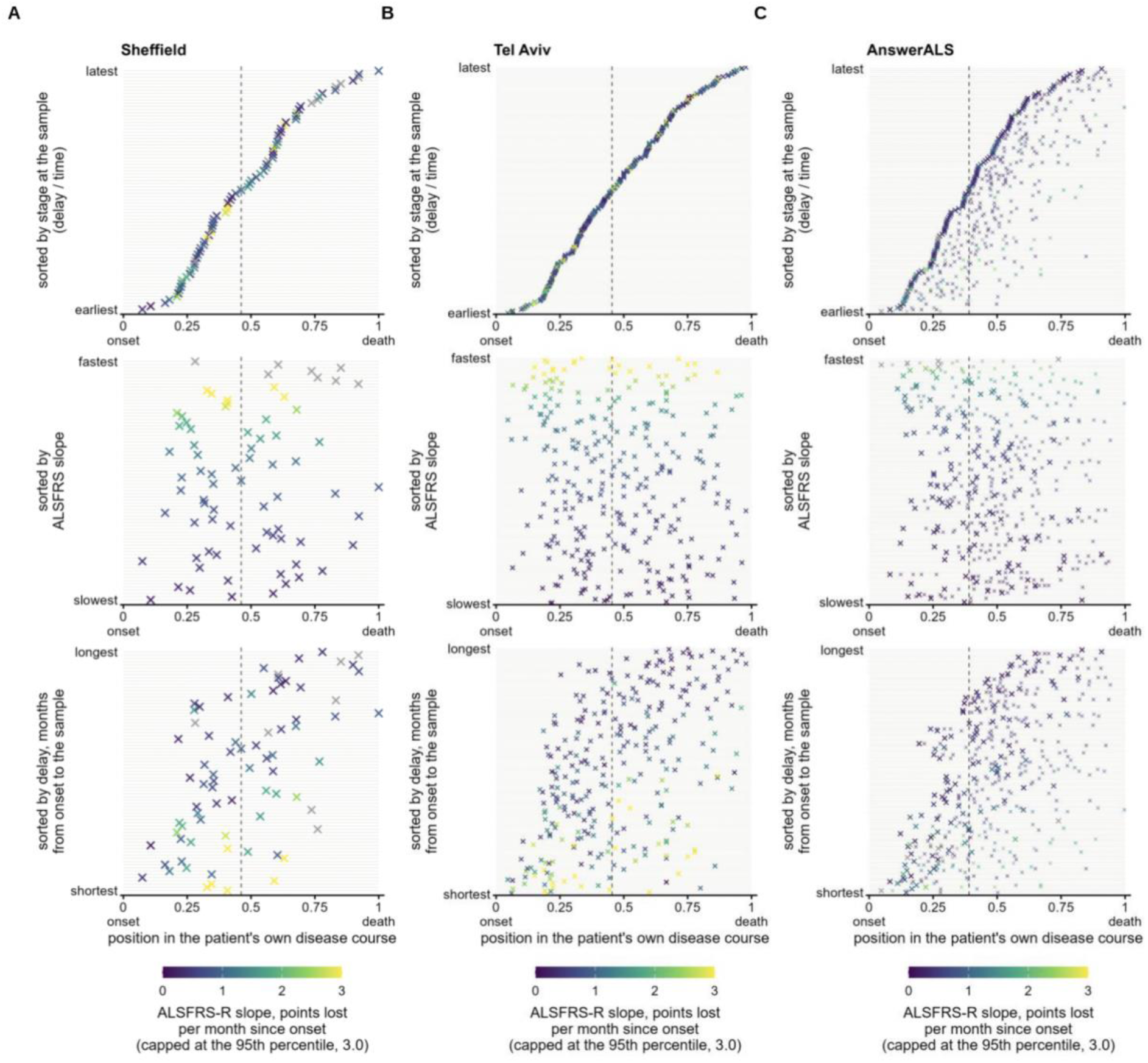
Disease stage at sampling in three ALS cohorts. Each row is one deceased patient with ALS, and each cross marks a sample at its disease stage: time from symptom onset to sampling divided by time from onset to death (0, onset; 1, death). Panels show (**A**) Sheffield (n = 75), (**B**) Tel Aviv (n = 290) and (**C**) AnswerALS (n = 129 patients with ALS; 431 samples). In AnswerALS, larger crosses are first samples and smaller crosses are later samples from the same patient, plotted on that patient’s row. Patients are ordered by disease stage at sampling (top), ALSFRS-R slope (middle) and months from onset to sampling (bottom). Colour shows ALSFRS-R points lost per month since onset, capped at the pooled 95th percentile (3.0); grey crosses mark patients without an ALSFRS-R slope. The dashed vertical line is the cohort median stage. ALS, amyotrophic lateral sclerosis; ALSFRS-R, revised ALS Functional Rating Scale.

**Supplementary Figure 3.**
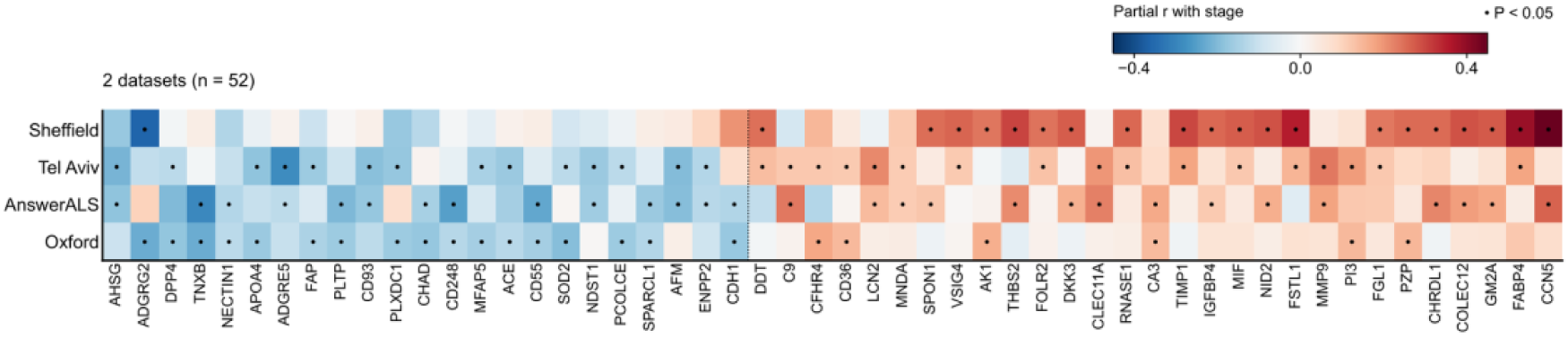
Stage-associated proteins significant in two datasets. Partial correlation with disease stage (adjusted for age and sex) of the 52 stage-associated proteins with P < 0.05 in exactly two of the four datasets, in Sheffield, Tel Aviv, AnswerALS and Oxford (rows); colour scale as in Figure 2H; dot, P < 0.05. The remaining 16 of the 68 proteins are shown in Figure 2H. Proteins are ordered lower (23) then higher (29) at later stage, then by mean stage effect.

**Supplementary Figure 4.**
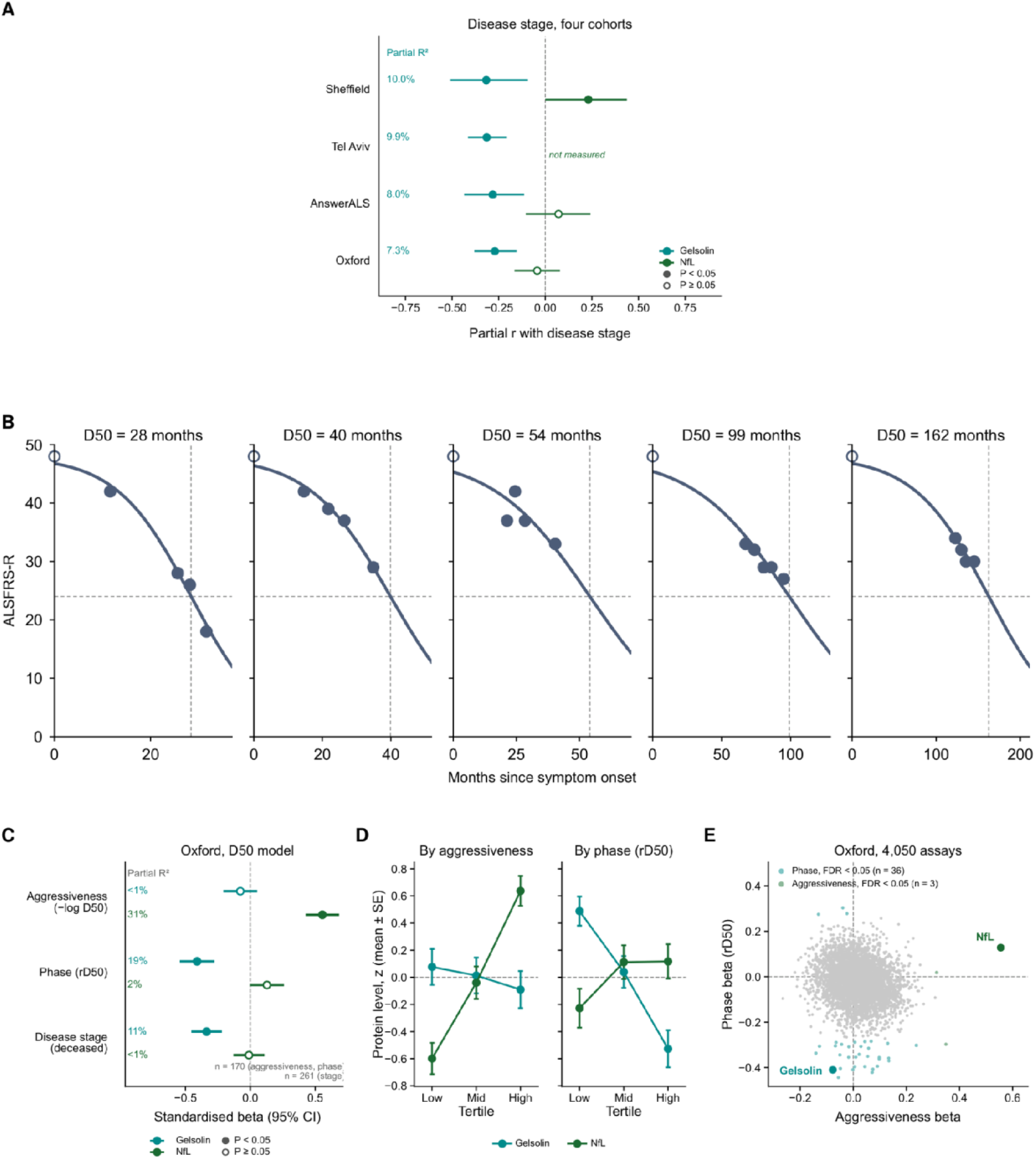
Gelsolin and NfL across cohorts and in the D50 model. (**A**) Partial correlation (95% CI), adjusted for age and sex, of GSN (dark cyan) and NfL (dark green) with disease stage in Sheffield (n = 75), Tel Aviv (n = 290), AnswerALS (n = 129) and the Oxford cohort (n = 260). Left, variance in stage explained by GSN (partial R²). NfL was not measured in Tel Aviv. (**B**) Examples of the D50 disease progression model: ALSFRS-R scores over time for five Oxford participants with at least four ALSFRS-R scores. They were chosen at the 5th, 25th, 50th, 75th and 95th percentiles of D50 among the 48 eligible participants. The D50 model^6^ fits a sigmoid curve (line) to each participant’s scores (filled circles), with a score of 48 assumed at symptom onset (open circle). D50 is the estimated time from onset to halving of the ALSFRS-R (score 24; horizontal dashed line), marked by the vertical dashed line and shown above each panel. Disease aggressiveness was defined as −log D50. Disease phase (rD50) at sampling was defined as time from onset to sampling divided by D50. (**C**) Oxford cohort, D50 model: standardised β (95% CI) of GSN and NfL for disease aggressiveness (−log D50) and disease phase (rD50), fitted jointly in 170 participants with at least two ALSFRS-R scores and adjusted for age, sex and study. The same panel shows disease stage in 261 deceased participants, with the same adjustment. Left, partial R² for each marker and measure. (**D**) Mean (± SE) z-scored GSN and NfL by tertile of aggressiveness and of phase in the 170 participants in C. (**E**) Aggressiveness β against phase β for 4,050 Olink assays in the Oxford cohort, from the models in C. Coloured points have FDR < 0.05 for phase (cyan) or aggressiveness (green), and GSN and NfL are labelled. ALS, amyotrophic lateral sclerosis; ALSFRS-R, revised ALS Functional Rating Scale; FDR, false discovery rate; GSN, gelsolin; NfL, neurofilament light chain.

**Supplementary Figure 5.**
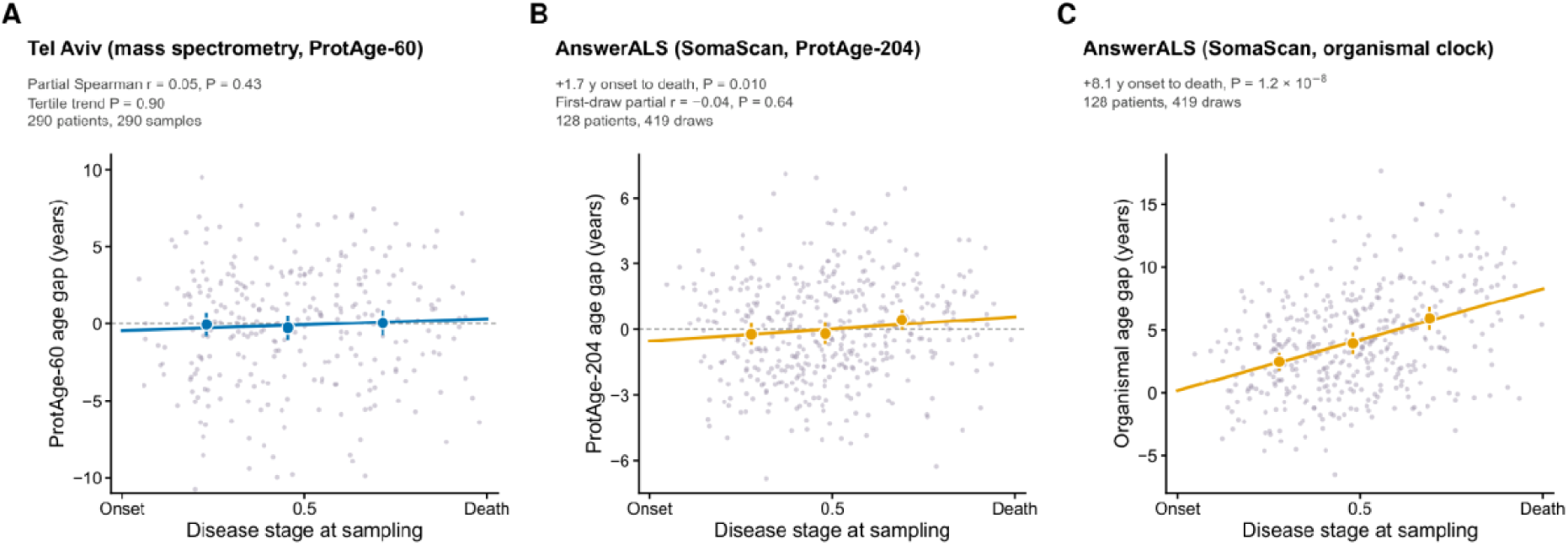
Proteomic age gap and disease stage on other proteomic platforms. (A) Tel Aviv, ProtAge retrained in UK Biobank on the 60 of 204 proteins measured by mass spectrometry (290 deceased patients with ALS). (B) AnswerALS, ProtAge-204 applied to SomaScan (162 of 204 proteins mapped; 128 deceased patients with ALS, 419 draws). (C) AnswerALS, organismal clock of Oh et al. (2023), trained on SomaScan (128 patients, 419 draws). Points show samples, large points stage-tertile means (95% confidence interval) and the line a linear fit. Disease stage is time from onset to sampling divided by time from onset to death. ALS, amyotrophic lateral sclerosis.^6^

## Supplementary Tables

**Supplementary Table 1.** Sheffield datasets and sample overlaps

**Supplementary Table 2.** Characteristics of the replication cohorts

**Supplementary Table 3.** Protein survival associations

**Supplementary Table 4.** Protein associations with disease stage

**Supplementary Table 5.** The 68 disease-stage proteins

**Supplementary Table 6.** Within-person change of circulating proteins in the Oxford cohort

**Supplementary Table 7.** Survival associations with and without adjustment for disease stage

**Supplementary Table 8.** Disease aggressiveness and phase (D50 model) in the Oxford cohort

**Supplementary Table 9.** Clinical determinants of the stage-associated proteins

**Supplementary Table 10.** Protein associations with age in UK Biobank and the ALS cohorts

**Supplementary Table 11.** Laboratory correlates of the protein and metabolite stage scores

**Supplementary Table 12.** Gene Ontology over-representation among stage-associated proteins: inflammation, immunity and muscle terms

**Supplementary Table 13.** Metabolite associations with disease stage and population ageing

**Supplementary Table 14.** Metabolon sub-pathway over-representation among stage-associated metabolites

## Notes

### Author Declarations

All data come from the Sheffield arm of the AMBRoSIA study (A Multicentre Biomarker Resource Strategy In ALS),28 (hereafter Sheffield). The London South East Research Ethics Committee (16/LO/2136) gave ethical approval for this work; the serum assayed here is stored under the Motor System Genomics approval (12/YH/0330). All participants gave written informed consent. People with motor neurone disease (MND; ALS, progressive muscular atrophy (PMA), primary lateral sclerosis (PLS) or progressive bulbar palsy (PBP)) were recruited between 6 June 2017 and 6 December 2023. Controls were volunteers without neurodegenerative disease.

